# Clonotype pattern in T-cell lymphomas map the cell of origin to immature lymphoid precursors

**DOI:** 10.1101/2021.07.05.21260052

**Authors:** Aishwarya Iyer, Dylan Hennessey, Robert Gniadecki

## Abstract

**Background:** Mature T-cell lymphomas (TCLs) are rare, clinically heterogeneous hematologic cancers of high medical need. TCLs have inferior prognosis compared with their B-cell counterparts, which is attributed to poor understanding of their pathogenesis. Based on phenotypic similarities between normal and neoplastic lymphocytes it has been assumed that TCLs develop in the periphery, directly from various subtypes of normal T-cells.

**Methods and findings:** To address the debated question of the cell of origin in TCLs we analyzed to identify the highly variable complementarity determining regions (CDR3) regions of T-cell receptor (TCR) to trace the clonal history of the T-cells. We have collected previously published whole genome-exome, and -transcriptome sequencing data from 574 TCL patients comprising five nodal lymphomas [anaplastic large cell lymphoma (n=67), peripheral T-cell lymphoma (PTCL, n=55), adult T-cell lymphoma/leukemia (n=135), natural killer T-cell lymphoma (NKCL, n=25), not specified/other (n=30)] and three extranodal, cutaneous T-cell lymphomas [mycosis fungoides (n=122), Sezary syndrome (n=130), and subcutaneous panniculitis-like T-cell lymphoma (n=10)]. TCR clonotypes contained in the tumor cell fraction, representing the clonotypes of malignant cells, were identified by de novo assembly of CDR3 regions of TCR γ, β and α. We have found that the vast majority of TCLs are clonotypically oligoclonal, although the pattern oligoclonality varied. Anaplastic large cell lymphoma was most diverse comprising multiple clonotypes of TCRγ, β and α whereas adult T-cell lymphoma/leukemia and peripheral T-cell lymphomas often showed monoclonality for TCRγ and β but had diverse TCRα clonotypes. These patterns of rearrangements were not compatible with the current mature T-cell precursor model and indicated that TCLs are initiated at the level of the lymphoid precursor. In keeping with this hypothesis, TCR rearrangements in TCLs resembled the pattern seen in the human thymus showing biased usage of V and J segments of high combinatorial probability resulting in recurrent, “public” CDR3 sequences shared between unrelated patients and across different clinical TCL entities. Frequencies of malignant clonotypes followed Zipf-Mandelbrot scaling law suggesting that TCLs comprise an interconnected system of expanding tumor clones.

The major limitation of this study is that it is based on the analysis of the TCR clonotypes and does not directly inform about developmental trajectories of cellular clones.

**Conclusions:** Lymphoid precursors are the likely cells of origin for mature T-cell lymphomas. Anaplastic large cell lymphoma seems to be derived from the most immature precursors with germline TCR whereas peripheral T-cell lymphoma and adult T-cell lymphoma/leukemia map to the later stages after TCRβ rearrangement stage. Clonotypically diverse initiating cells may seed target tissues being responsible for disease relapses after therapy.

## Introduction

T-cell lymphomas (TCLs) is a heterogenous group of 29 malignancies comprising approximately 10% of non-Hodgkin’s lymphomas and the incidence of 6000 cases per million [1,2]. Most TCLs are classified as mature T-cell lymphomas because malignant cells phenotypically resemble mature T-cells, harbor rearranged T-cell receptor (TCR) V(D)J genes and many express TCR αβ heterodimer [3,4]. Striking similarities between subsets of normal T-cells and the malignant cells in TCLs (e.g. to T-regulatory cells in the adult T-cell lymphoma/leukemia or central memory T-cells to Sezary syndrome) led to the general acceptance of the theory that different subsets of mature T-cells are the cells of origin in mature lymphomas [5].

TCR gene sequences are excellent markers of T-cell lineage, because TCRγ, TCRβ, and TCRα loci are sequentially rearranged during different stages of intrathymic maturation of T cells from diverse pool of variable (V), diversity (D), and joining (J) gene. The TCR β locus contains 47 V (*TRBV*), 2 D (*TRBD*) and 13 J (*TRBJ*) segments whereas the TCR α locus comprises 42 *TRAV* and 61 *TRAJ* segments, which recombine yielding unique DNA sequences that are retained in genomes of all daughter cells [6]. Analysis of the complementarity determining region 3 (CDR3), the most diverse fragment of TCR chain coded by the V-(D)-J joining region and involved in antigen recognition, has long been used in the molecular diagnosis of T-cell lymphomas and leukemias. The repertoire of normal human TCR is in the range of 10^6^-10^7^ clonotypes (unique CDR3 sequences defining a T-cell clone) [7–9] whereas in mature TCLs it is dominated by a single clonotype, representing the clonally expanded malignant cells (i.e. malignant clonotypes) [10].

Clonotypic analyzes of T-cell lymphomas and leukemias are not only useful for diagnostic purposes but might also provide important clues to the pathogenesis. Transformation of the mature T-cell would result in perfect monoclonality in all TCR loci because TCR rearrangement does not occur in mature T-cells and TCRα and TCRβ clonotypes would be inherited from the cell of origin. Transformation during earlier stages of T-cell development, before the completion of the final TCRα rearrangement, would produce oligoclonal or polyclonal patterns of clonotypes, characteristic for the stage in which the transformation happened. Indeed, our previous analyses of malignant T-cell clonotypes in cutaneous T-cell lymphoma revealed an oligoclonal pattern of TCRβ and TCRα, but monoclonality of TCRγ suggesting that original malignant transformation occurred at the stage of a double-negative thymocyte after rearrangement of the TCRγ locus [11,12]. A similar pathogenic scenario involving early thymocytes has been proposed for the anaplastic large cell lymphoma [13,14], but it is unclear whether other TCLs might also be derived from immature T-cells.

During analysis of our CTCL sequencing data we were also puzzled by the finding that certain malignant clonotypes were repeatedly found in samples from unrelated patients whereas due to the vastness of TCR repertoire we had expected that malignant clonotypes would have been unique for each patient. We have noted a preferential usage of certain Vβ and Vα segments in cutaneous T-cell lymphomas [11] and were intrigued by the finding that similar biases in V segment usage was reported in T-cell acute lymphoblastic leukemia (T-ALL) which is derived from lymphoid precursors arrested at various stages of development [15,16].

In this study we employed the previously optimized bioinformatic pipeline for de novo assembly of CDR3 regions using paired-end RNA sequencing, whole genome- and whole exome DNA sequencing [11] to analyze malignant TCR clonotypes in 574 samples comprising 8 major subtypes of TCL. We propose that TCR rearrangement pattern across different clinical TCL entities is incompatible with the mature T-cell transformation hypothesis and supports the immature lymphoid progenitor as the cell of origin.

## Methods

### TCL data collection and TCF analysis

The graphical summary of the methods is shown in **Fig 1**. Raw fastq files of whole exome, whole genome and whole transcriptome sequencing and whole transcriptome sequencing were collected from 13 different studies (574 patients)[17–29]. To access the human genome data from restricted databases such as dbGaP and EGA was provided by Health Research Ethics Board of Alberta Cancer Committee under ethics ID-HREBA.CC-20-0118. Additionally 50 healthy controls samples were also obtained[30]. The accession number of the studies are listed in **supplementary Table S1** and the diagnostic grouping in the **supplementary Table S2**. Tumor cell fraction (TCF) was determined based on copy number aberrations (CNA) using Titan CNA(v.1.17.1) [31] and/or Ichor CNA (v. 0.2.0) [32] in the 368 samples for which DNA sequencing data were available **(supplementary Table S3)**. Dataset of Crescenzo et al. (SRP044708) lacked control DNA and therefore, a panel of normal as recommended in the GATK pipeline [33] was created to analyze the TCF for samples from that study. Majority of transcriptome data was not obtained from the same samples as their exome counterparts and therefore, TCF data calculated using the DNA sequencing was not applicable to the TCR identified from the RNA sequencing.

**Figure 1.**
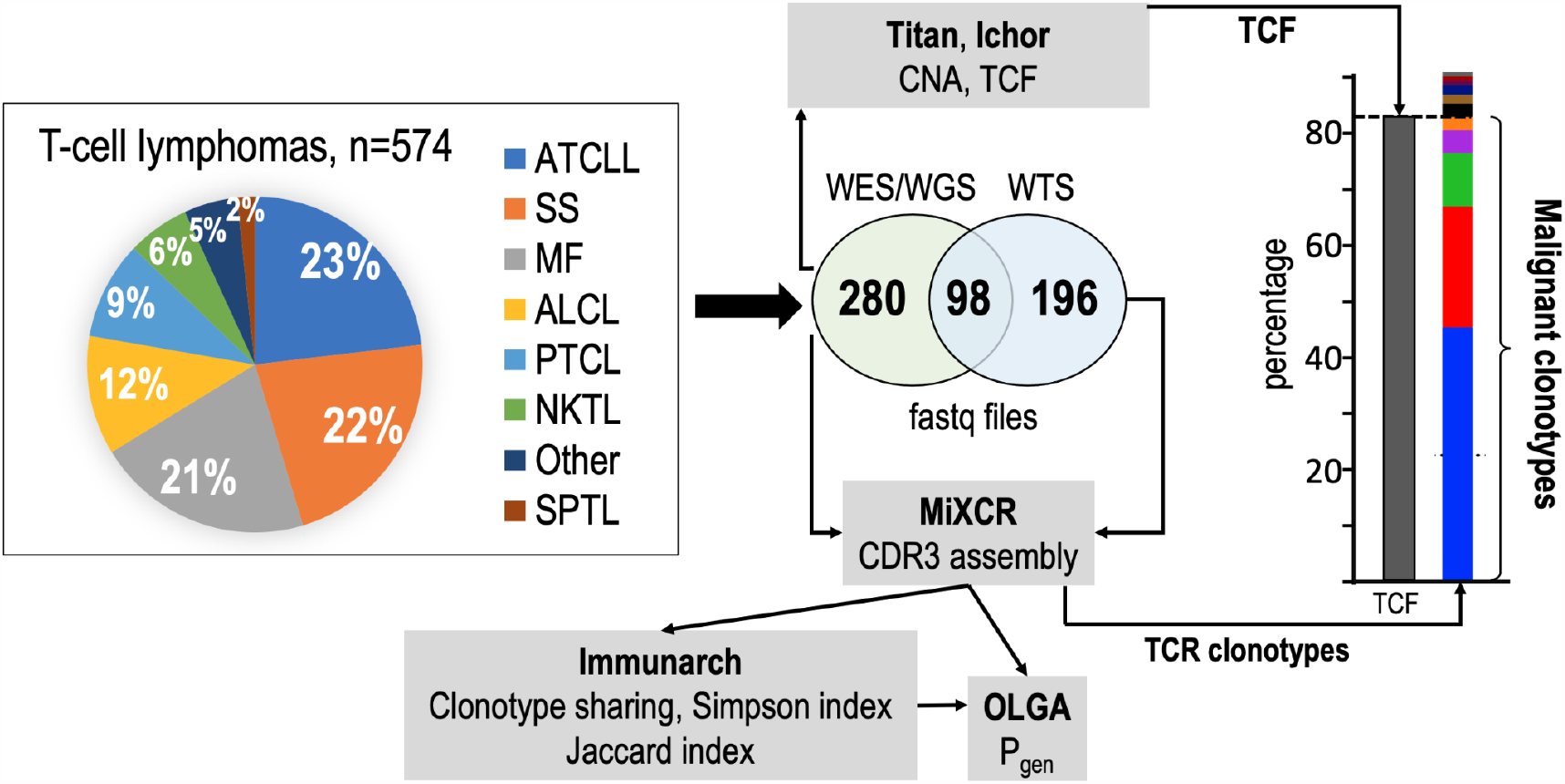
Summary of the methodological approach to TCR analysis in TCLs. Pie-chart representing the percentage of sequencing samples collected from different subtypes of TCLs were analyzed for tumor cell fraction (TCF) and the TCR clonotypes. The diagnostic groupings are detailed in supplementary Table S2. TCF was estimated based on the chromosomal aberration identified using TitanCNA or IchorCNA. TCR clonotypes were identified using MiXCR and the diversity index (inverse Simpson), clonotype sharing (Jaccard index), and probability of clonotype generation (P_gen_) were analyzed using Immunarch and OLGA pipelines, as detailed in the Methods. WES, whole exome sequencing; WTS, whole transcriptome sequencing; WGS, whole genome sequencing; ATCLL, adult T-cell lymphoma/leukemia; SS, Sezary syndrome; MF, mycosis fungoides; ALCL, anaplastic large cell lymphoma; PTCL, peripheral T-cell lymphoma; NKTL, natural killer/T-cell lymphoma; SPTL, subcutaneous panniculitic T-cell lymphoma.

### TCR clonotype identification and quantification

TCR clonotypes were identified and quantified using MiXCR version 3.0.3 or 3.1.0 [34]. We used the superscript to indicate whether the clonotype was identified by the DNA, RNA, or amino acid (aa) sequence (e.g. TCRα^DNA^). Default recommendations for analysis of exome and transcriptome data were used with the exception of eliminating assembly of short reads to identify additional TCR clonotypes. Short and long read alignments were included for whole-transcriptome sequencing (WTS); however, for WES data, partial reads were filtered out because they might be the captures of only V or J sequences. After identification of clonotypes further elimination of clonotypes based on mutational positions were performed and best predictive VJ combinations were included in further analyses. For the cases where TCF was available, the malignant clonotypes (i.e. clonotypes of the malignant T-cells) were determined by including the most abundant clonotypes to match TCF of the sample **(Fig 1)**. Shared clonotype identification was based on TCRβ^aa^ and was used to calculate diversity (inverse Simpson index) and clonotype sharing (Jaccard index) with Immunarch version 0.6.6 [35]. Probability of generation of CDR3 (P_gen_) was calculated using OLGA version 1.2.3[36] for the TCL samples and the 50 healthy controls. Immunarch, Graphpad Prism (version 9) and Excel (version 16.16.27) were used for data representation.

## Results

### Clonotypic TCRβ diversity in T-cell lymphomas

We analyzed data from 574 samples comprising the following TCL diagnoses: mycosis fungoides (MF, a primary cutaneous T-cell lymphoma, n=122), Sezary syndrome (SS, leukemic form of primary cutaneous T-cell lymphoma, n=130), anaplastic large cell lymphoma (ALCL, n=67), adult T-cell lymphoma/leukemia (ATCLL, n=135), natural killer T-cell lymphoma (NKTL, n=25), peripheral T-cell lymphoma (PTCL, n=55), subcutaneous panniculitis-like T-cell lymphoma (SPCL, n=10). We were unable to classify 30 samples into one of the above categories, which were grouped as “other TCLs” (**supplementary Table S2**).

To determine the sequences of CDR3, we used the previously described bioinformatic analysis of bulk sequencing data from whole exome sequencing (WES), whole genome sequencing (WGS), and whole transcriptome sequencing (WTS) [11]. To differentiate between malignant clonotypes (i.e. clonotypes derived from malignant T-cells) and those contributed from the infiltrating reactive cells, we ranked the TCR^DNA^ clonotypes (clonotypes obtained from DNA sequencing) starting with the most abundant and included only those of the cumulative frequency matching the tumor cell fraction (TCF) in the sample **(Fig 1)** [12].

Among 378 samples for which genomic data (WES/WGS) was available to calculate TCF, the majority (75%) showed more than one TCRβ^DNA^ clonotype attributed to malignant T-cells (matching TCF) **(Fig 2**). We considered three rearrangements as evidence of oligoclonality because although approximately 60% of T-cells rearrange TCRβ on only one chromosome, the remaining 40% are bi-allelic rearrangements (the 60/40 rule) [37,38]. Oligoclonality of malignant clonotypes was most prevalent in ALCL (43/47) and least common in NKTL (12/25) (**Fig 2**).

**Figure 2.**
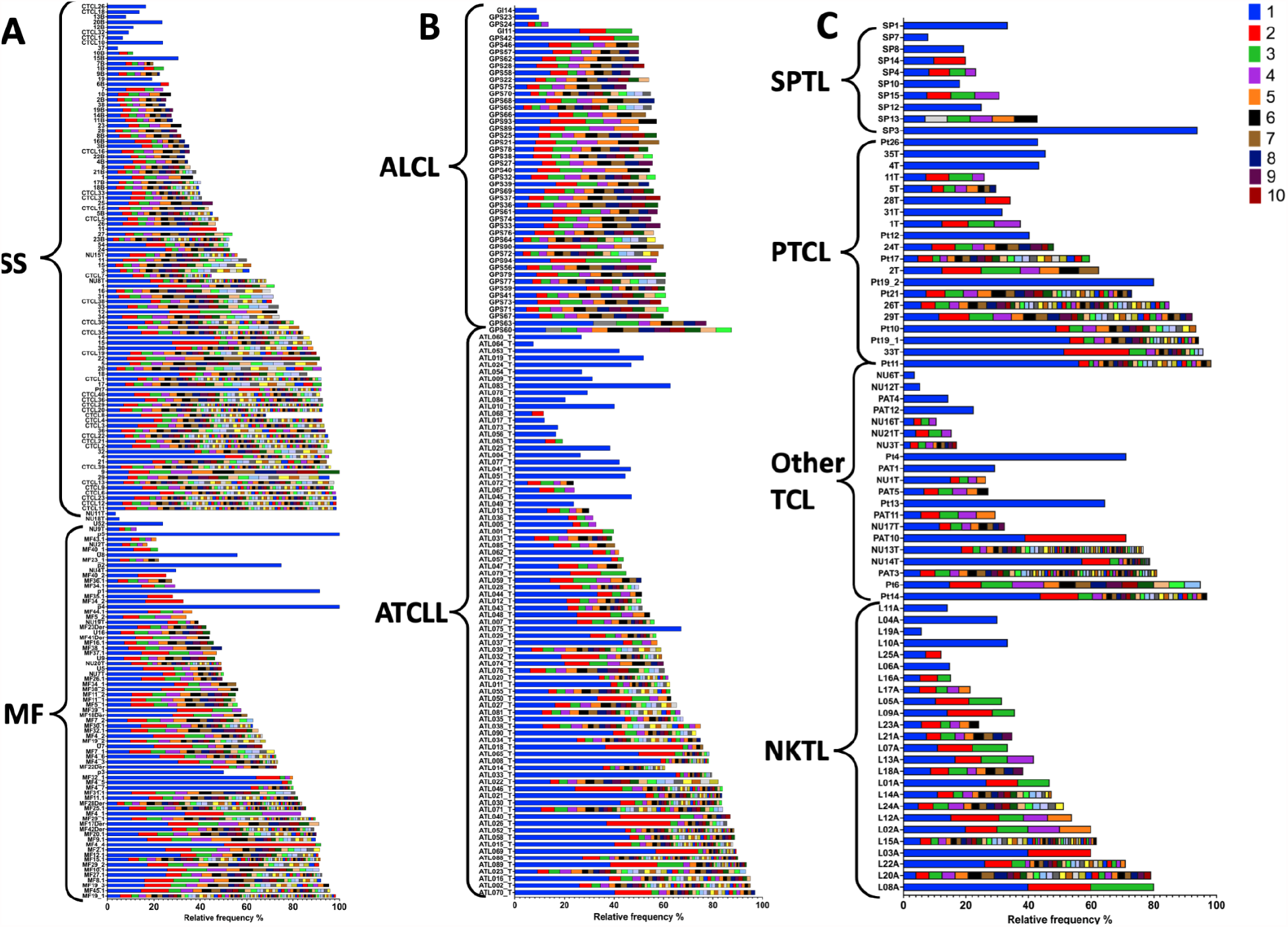
Frequency of TCRβ^DNA^ clonotypes in TCLs. Total of 378 samples from 8 subtypes of T-cell lymphomas (TCLs) were analyzed for identifying tumor cell fraction (TCF) and TCRβ^DNA^ clonotypes using the whole exome/genome sequencing data. TCRβ^DNA^ clonotypes corresponding to the TCF are indicated by the coloured bars in the descending order of relative frequencies (the most abundant, rank 1, is the dark blue bar). The data are split by diagnosis: **A** - cutaneous lymphomas, MF and SS; **B** - ATCLL and ALCL, **C** - NKTL, PTCL, SPTL and Other. Abbreviations of disease entities as in Fig 1.

The allelic exclusion of TCRβ locus assures that a mature T-cell only expresses one clonotype [37], even in the presence of biallelic rearrangements [39,40]. Thus, the single TCRβ clonotype at the level of RNA (TCRβ^RNA^) unequivocally identifies the T-cell clone. Since TCF could not be calculated for WTS samples because the matching WES/WGS data were not available, we analyzed TCRβ clonality by applying the relative frequency threshold of 25%. This threshold has been proposed by other investigators [41,42] and we considered it reasonable to adopt because the majority (79% of TCL samples) had TCF of 25% or higher **(supplementary Table S3)**. In 26% of samples the most abundant TCRβ^RNA^ clonotype had a frequency below 25%, i.e. the criterion of monoclonality was not met (**Fig 3**). In 6 samples the relative frequency of the second most frequent TCRβ^RNA^ clonotype was >25% and in 38 cases the frequencies of the first and the second clonotypes were comparable (no single dominant clonotype), indicating oligoclonality of malignant clonotypes.

**Figure 3.**
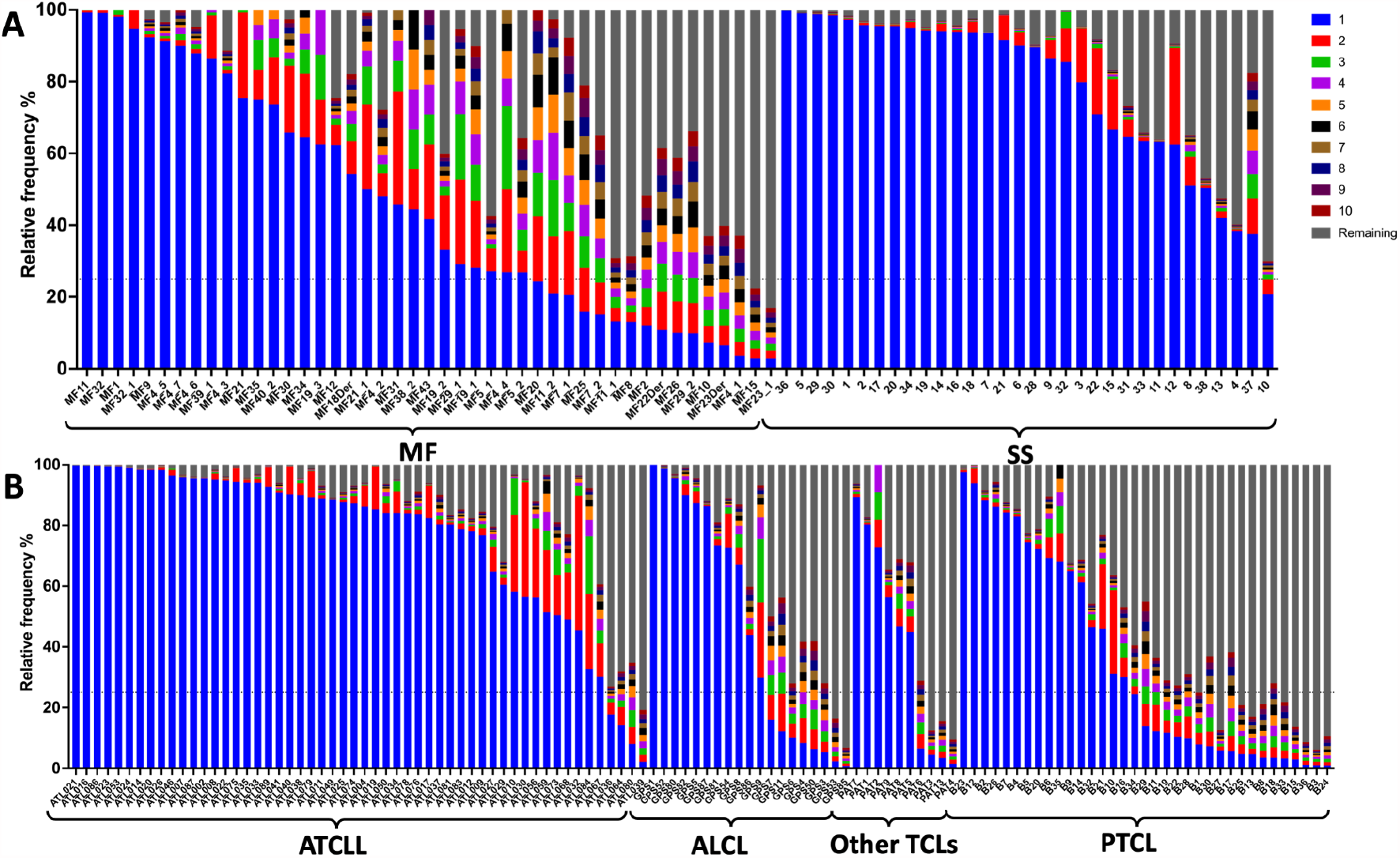
Frequency of TCRβ^RNA^ clonotypes in TCLs. Total of 196 RNA samples from 6 subtypes of TCLs were analyzed to identify the frequency of the TCRβ^RNA^ clonotypes. Ten most abundant clonotypes are indicated in colour using the same coding as in Fig 2; the remaining clonotypes are merged and their combined frequency indicated by the gray bar. The data are split by diagnosis: **A** - cutaneous lymphomas, MF and SS; **B** - ATCLL, ALCL, PTCL and Other. Abbreviations of disease entities as in Fig 1.

To further assess the diversity of the TCRβ clonotypes using the inverse Simpson index which varies from 1 (one clonotype) to the maximum value representing the number of clonotypes (in our analysis n=10), taking into account clonotype concentration **(Fig 4)**. For the TCRβ^DNA^ clonotypes, the diversity was highest in SS and ALCL and lowest in ATCLL. The diversity of TCRβ^RNA^ clonotypes had a slightly different pattern with a decrease in diversity for SS and ALCL when compared to TCRβ ^DNA^ clonotypes. This can be explained by the increase in the relative contribution of the most abundant TCRβ^RNA^ clonotype (higher clonotype concentration) relative to TCRβ^DNA^ which, in turn, is likely to be caused by a higher proportion of non-productively rearranged TCRβ loci. The mean diversity for MF, ATCLL, and PTCL remain the same for TCRβ^DNA^ and TCRβ^RNA^.

**Figure 4.**
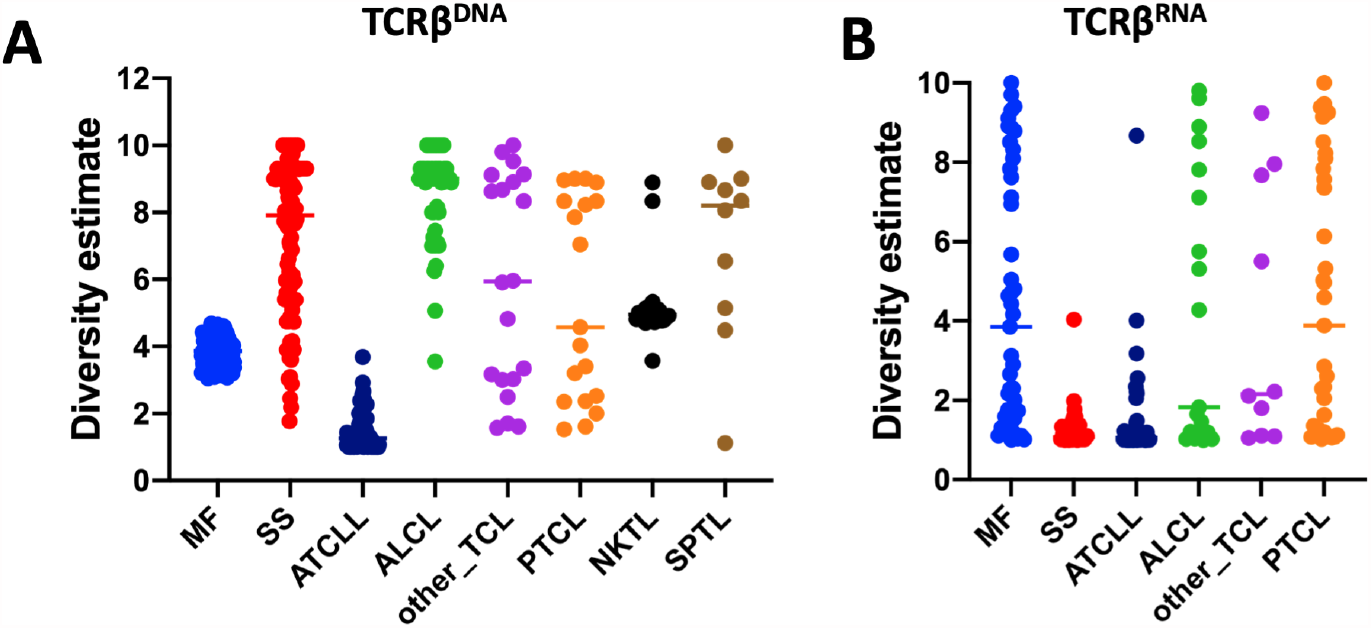
Diversity of TCRβ clonotypes in different TCLs. Inverse-Simpson index was calculated for the ten most abundant TCRβ^DNA^ **(A)** and TCRβ^RNA^ clonotypes **(B)** and individual samples presented in the dot-plot graph. Abbreviations of disease entities as in Fig 1.

### Patterns of TCRγ, -β and -α corroborate a branched evolution model of TCLs

The analyses described above suggested that TCR monoclonality is not a constant feature of TCLs and approximately ⅓ of all samples display an oligoclonal pattern of malignant TCRβ, which is incompatible with the hypothesis of the mature T-cell origin hypothesis. To explore this further, we analyzed the frequencies of TCRγ and -α clonotypes. Unlike TCRβ, TCR-γ and -α loci are not subjected to allelic exclusion and are usually rearranged at both chromosomes **(Fig 5 A)**. Moreover, TCRγ is rearranged before TCRβ so the cells with the same TCRγ rearrangement may harbor several TCRβ rearrangements. The same is true for TCRα; a single TCRβ clone may comprise several (usually 2-4) TCRα clonotypes because of biallelic rearrangement and secondary rearrangements during expansion of the TCRβ clone **(Fig 5 A)**. It is thus possible to deduce the stage of T-cell transformation from the relative frequencies of TCR γ, β and α.

**Figure 5.**
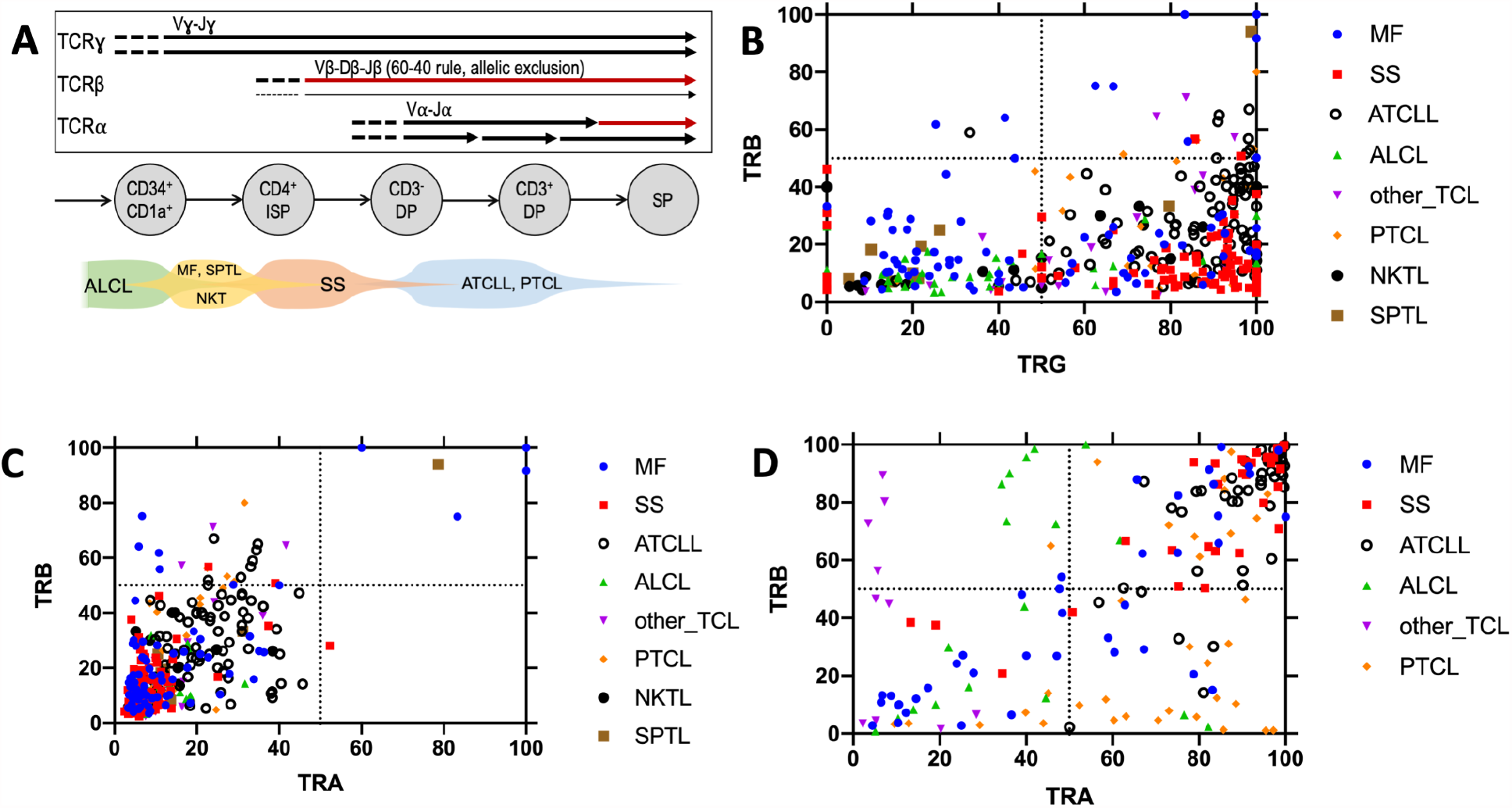
Oligoclonality in T-cell lymphomas. **A** - Schematic presentation of the TCR gene rearrangement during T-cell development. The arrows represent rearrangement on each chromosome mapped to different stages of normal human thymocyte. The thin arrowline for TCRβ symbolizes the 60-40 rule. Predicted time points of the transformation in various subtypes of TCLs is shown in the bottom row. **B, C** - WES/WGS samples (n=367) were analyzed to identify the TCRα^DNA^, TCRβ^DNA^ and TCRγ^DNA^ clonotypes. The frequency of the most abundant TCRβ^DNA^ clonotype was plotted versus the added frequency of two most frequent TCRγ^DNA^ clonotypes or the two most abundant TCRα^DNA^ clonotypes **(C)** for each sample. **D** - A similar correlation plot of the frequency of dominant TCRβ^RNA^ clonotype versus the dominant TCRα^RNA^ clonotype (data from 196 WTS samples). Abbreviations of disease entities as in Fig 1.

Interestingly, most TCL samples showed a clearly monoclonal pattern of TCRγ dominated by one or two clonotypes **(Supplementary Fig S1)**. By plotting the frequency of the dominant (most abundant) TCRβ^DNA^ clonotype versus that of the two most abundant TCRɣ clonotypes (**Figure 5 B)** most of the cases fall into two categories. The samples located in the lower left quadrant had low-frequency TCRβ^DNA^ and low-frequency TCRγ and represented 32% of samples classified as oligoclonal for both TCRγ and TCRβ. Most ALCL cases were found in this group. The remaining (68%) of the samples showed a high proportion of TCRγ and a disproportionately low frequency of TCRβ^DNA^ (lower right quadrant) indicating TCRγ monoclonality but oligoclonality with respect to TCRβ. Only single cases of TCL were in the right upper quadrant representing high frequency of TCRβ and TCRγ which would be characteristic for an expansion of a single, mature T-cell clone. A similar plot of the frequencies of the dominant TCRβ^DNA^ vs the combined frequencies of the first and second (biallelic) TCRα^DNA^ showed a linear correlation within the lower left quadrant representing oligoclonality in both chains (**Fig 5 C**). The samples clustered in two major groups: those with the higher frequency of TCR β and α (mostly ATCL) and the remaining occupying the low-frequency portion of the quadrant (**Fig 5 C**). The analysis of the TCRα^RNA^ and TCRβ^RNA^ clonotypes helped in the interpretation of the findings. As shown in **Fig 5 D**, most ATCLL and SS samples showed monoclonality being located in the right upper quadrant representing high frequencies of the dominant clonotypes. However, ALCL, PTCL, MF and other TCL were scattered in the remaining quadrants underscoring their oligoclonality not only at the DNA level but also at the level of TCR gene expression.

Although the above described findings showed significant variation in TCR clonality patterns within the same TCL diagnostic entities, general conclusions can be made regarding the putative cell of origin **(Fig 5 A)**. ALCL was the most heterogeneous with the clear oligoclonality of TCRγ, -β, and -α which indicated that the transformation takes place in immature thymocytes before TCRγ recombination step. On the opposite pole of the spectrum were SS and ATCLL which in the majority of cases showed TCRγ monoclonality and high frequency of TCRβ, mapping the transformation event approximately at the TCR β rearrangement step. However, none of the TCLs exhibited a consistent pattern of monoclonality, defined at the DNA level by maximum two rearrangements of each TCRγ, -β and -β genes matching the TCF and at the RNA level by a single dominant TCRβ clonotype and maximum two TCRα clonotypes.

Based on the above described results we hypothesized that TCRβ^DNA^ clonotypes may label neoplastic T-cell clones in the sample and the distribution of those clonotypes is not random but might be a result of the clonal evolution of the disease. Systems comprising objects which grow in size and compete with each other (such as growth of cancer subclones in a tumor) often obey power laws such as Zipf-Mandelbrot law expressed as: *S*_(*k*)_ ∼ *S*_(1)_ /*k*^γ^ where *S*_*(k)*_ is the size of the species (in our case, the size of the TCRβ^DNA^ clonotype) of the rank *k, S*_*(1)*_ is the size of the first ranked species and γ is a scaling constant [43,44]. Indeed, the log-log plots in which mean abundance of the TCRβ^DNA^ clonotype was plotted versus the its rank revealed the expected linear relationship for all TCLs, although the slope of the linear regression line varied from −0.34 for ALCL to −1.21 for ATCLL reflecting different values of the constant γ for different types of TCLs **(Fig 6)**. Reproducible linearity of the rank-size TCRβ plots for different TCLs were therefore consistent with zipfian dynamics [45] and supported the hypothesis that malignant T-cell clones are dynamically coherent and interactively evolve via neutral clonal evolution [44], as postulated by us for MF [17].

**Figure 6.**
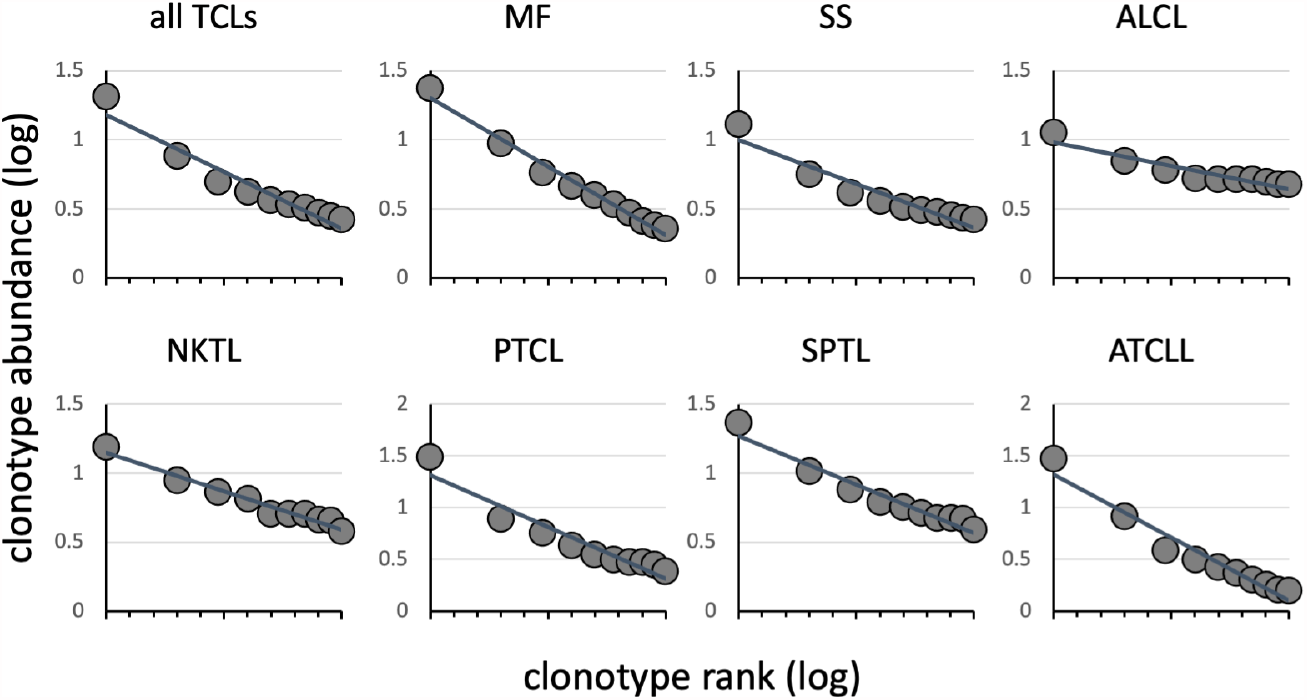
Rank-frequency plots showing power-law distribution of clonotypes in TCLs. TCRβ^DNA^ clonotypes identified from 378 TCL samples were analyzed to estimate the frequency of each clonotype in the individual samples. The frequency of each clonotype was plotted versus its abundance rank (log-log plot). **A** - TCRβ^DNA^ clonotypes from all TCLs subtypes. **B-H -** TCRβ^DNA^ clonotypes from individual TCL entities. The line is the linear regression. Abbreviations of disease entities as in Fig 1.

### Biased V and J usage in TCLs

Previous studies have shown biased rearrangement of certain TCR V and J genes combinations [46,47], but it is not clear whether those findings are generalizable to all TCLs. We calculated the frequencies with which V and J genes are represented in the most abundant clonotype of TCRβ **(Fig 7)** or the two most abundant clonotypes in case of TCRα and TCRγ **(Fig 8)**. There was a strong preferential usage of certain Vβ and Vα segments across different TCLs. The most striking was Vβ20-1 (*TRBV20-1*) which was found in a large proportion of clonotypes in MF, SS, ATCLL, ALCL and NKTL, both at the level of DNA and RNA. Vβ4-1 (*TRBV4-1*) was overrepresented in ALCL, NKTL, and SPTL. Other Vβ segments were more disease-specific, e.g. *TRBV18* was often found in MF and *TRBV24-1* was frequent in PTCL. Biased use of Vα segments was also obvious, for example Vα16 (*TRAV16)*, Vα35 (*TRAV35)*, Vα41 *(TRAV41)* and Vα3 (*TRAV3)* were overrepresented in most TCLs. The Jβ and Jα segments also seemed to be unequally used (e.g. *TRBJ2-1* and *TRBJ2-7* overexpression in all subtypes of TCLs) but because of a small number of J genes the patterns are less obvious (**Fig 7 and 8**).

**Figure 7.**
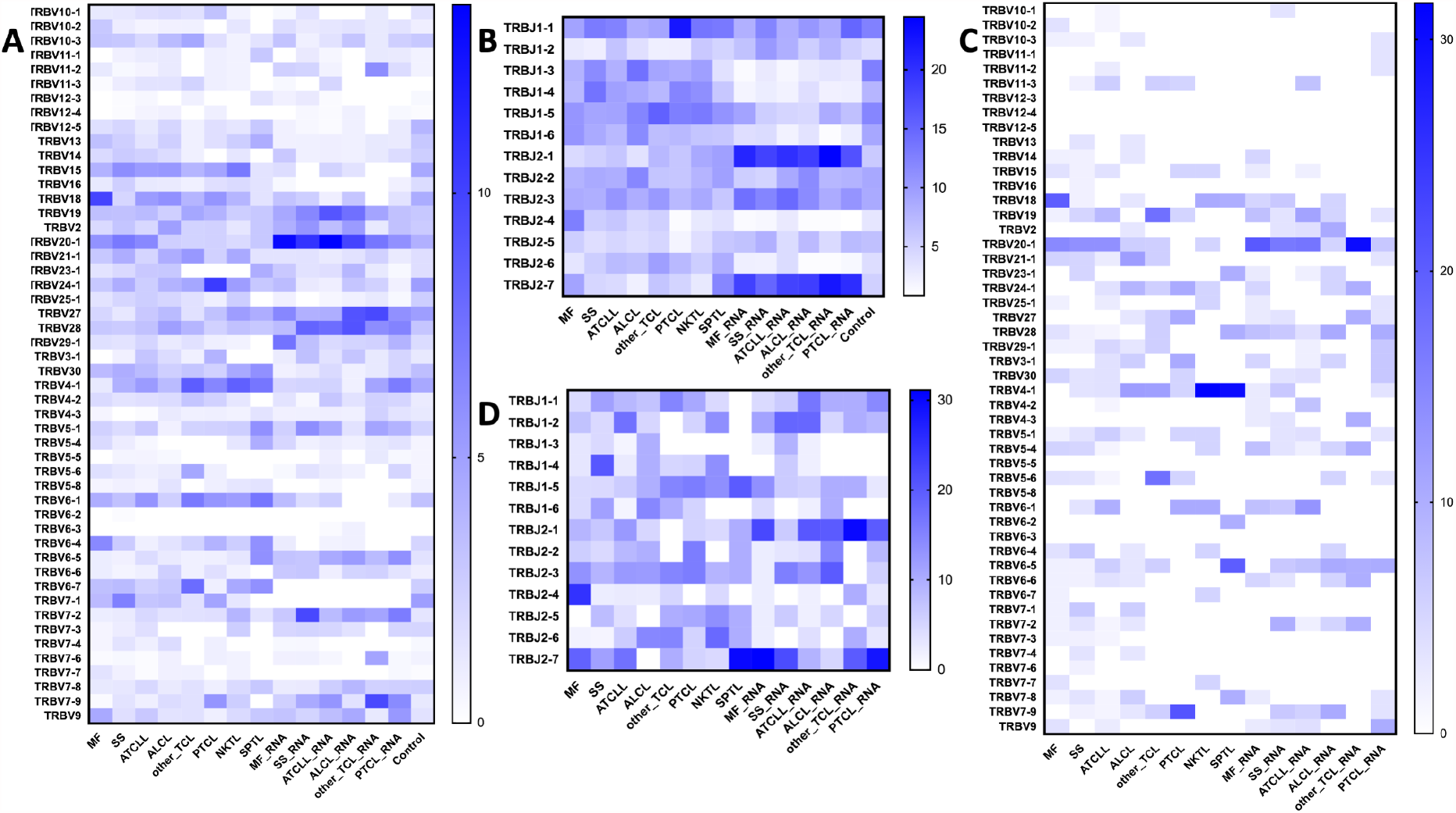
Vβ and Jβ (*TRBV/TRBJ*) usage in TCLs. The identified TCRβ^DNA^ and TCRβ^RNA^ clonotypes from WES/WGS and WTS data for the TCL diagnoses and control blood lymphocytes to identify preferential *TRBV* and *TRBJ* segment usage. **A, B** - represent *TRBV* and *TRBJ* frequencies, respectively, in the 10 most abundant clonotypes. **C, D** - similar plot as in A, B but including the single, most abundant clonotype. Control group is absent in C and D due to the lack of a high frequency clonotype in these samples. Abbreviations of disease entities as in Fig 1.

**Figure 8.**
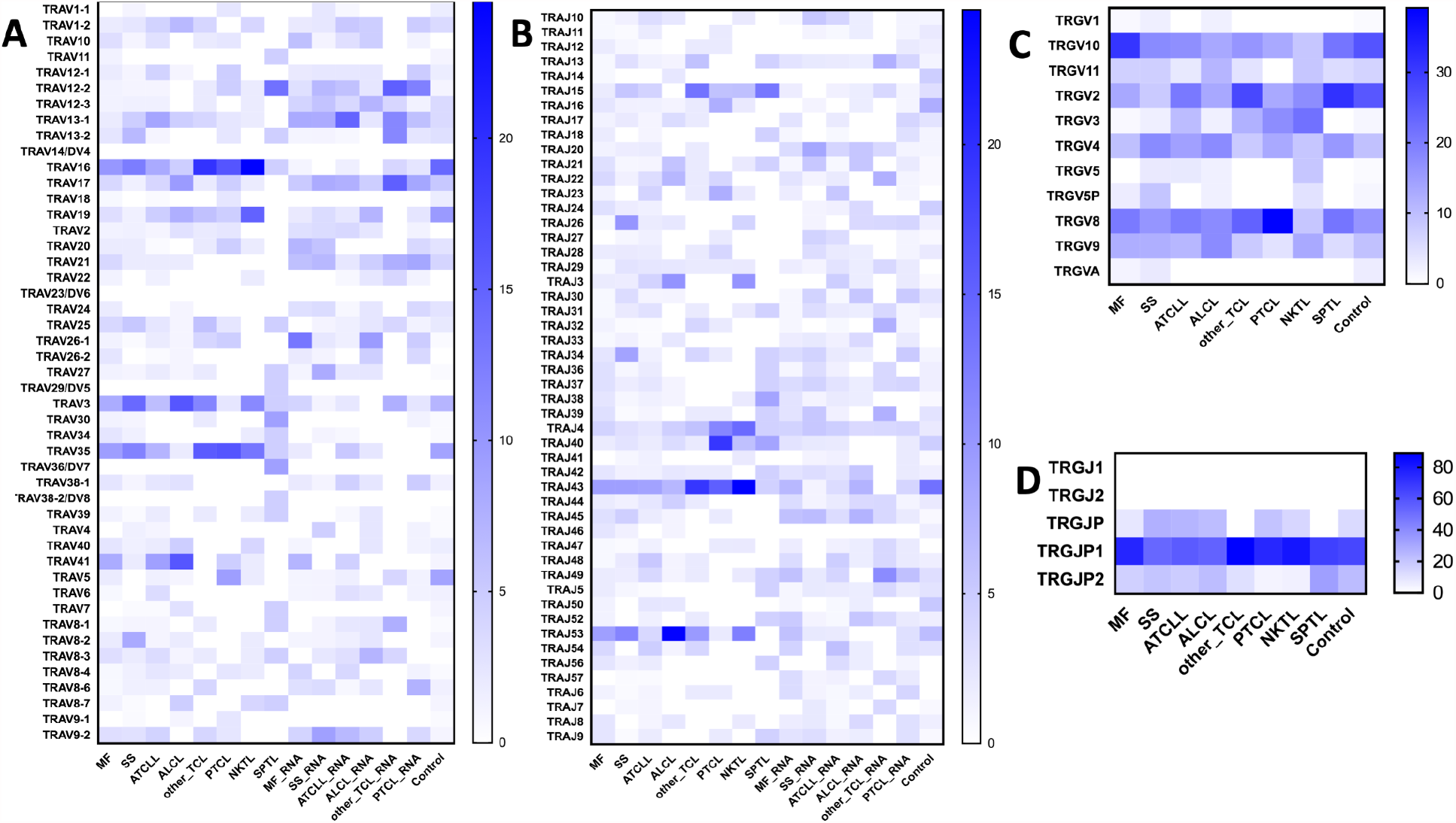
V and J (*TRAV, TRAJ, TRGV, TRGJ*) usage in TCLs. The frequency of V and J segments of TCRα and TCRγ clonotypes are plotted as in Fig 7. Two most abundant TCRα^DNA^ TCRγ^DNA^ clonotypes from TCLs and control samples were identified and the frequency of V and J segments plotted on the heat map. The RNA samples were used only for identification of the TCRα^RNA^ clonotypes. **A, B** - TRAV and TRAJ segment usage; **C, D** - *TRGV* and *TRGJ* segment usage.

We have also analyzed V and J segment usage in the 10 most abundant TCRβ clonotypes, representing the median number of malignant clonotypes, determined in comparison to TCF. The overall pattern was similar to that of the most abundant clonotypes confirming biased use of *TRBV20-1, TRBV4-1, TRBJ2-1* and *TRBJ2-7*.

Lastly, previous studies in CTCL have indicated over-representation of the *TRGV10-TRGJP1* in the malignant T-cells [48]. We could confirm this in our analysis and also found that *TRGV2, TRGV8* and *TRGV9* were frequently used across different TCLs, but *TRGV1, 5, 11* were relatively underrepresented (**Fig 8 C,D**).

### Shared malignant clonotypes represent public clonotypes and have high recombination probability

The results described above indicated that certain V-J combinations are overrepresented suggesting that the clonotypes (defined as by the aminoacid sequence of the highly variable CDR3 region) may also be shared between patients with different TCLs. Indeed, as shown in **supplementary Tables S4 and S5** we have found numerous shared TCRα^aa^ and TCRβ^aa^. Some clonotypes were shared across a large number of patients with different TCLs. For example, CARRKSSFF was detected in 100 samples (54 samples using *TRBV28-TRAJ1-1*) and CDNNNDMRF (*TRAV16-TRAJ43*) was present in 213 samples.

We estimated the fraction of the malignant clonotypes shared by two or more individuals using the Jaccard index [ranging from 0 (no overlap) to 1 (full overlap of TCR repertoires). Both TCRα^aa^ and TCRβ^aa^ had a high degree of sharing (Jaccard indices of 0.4 and 0.34, respectively), predictably showing higher sharing for TCRα.

Shared malignant clonotypes resembled public clonotypes found in the blood of healthy subjects. Public TCRs largely result from V-J recombination biases and convergent recombination where rearrangement of different segments can result in the same aminoacid sequence [49]. Public TCR chains can pair with a diverse repertory of “private” TCR chains and therefore they are not determining antigen specificity [49]. To determine whether the detected, shared malignant clonotypes might represent the result of biased, convergent V-J recombination, we calculated the probability of CDR3 sequence generation (P_gen_) value [36,50] for the most abundant TCRβ and TCRα clonotypes. P_gen_ of those shared malignant clonotypes had two probability peaks at 10^−9^ and 10^−6^ respectively (**Fig 9 A** and **supplementary Tables S4-S7**). Shared TCRβ^RNA^ and TCRα^RNA^ had even higher values with the maxima at 10^−6^ and 10^−4^, respectively (**Fig 9 B**) which were significantly higher than the Pgen of peripheral blood private (unique) clonotypes (<10^−9^).

**Figure 9.**
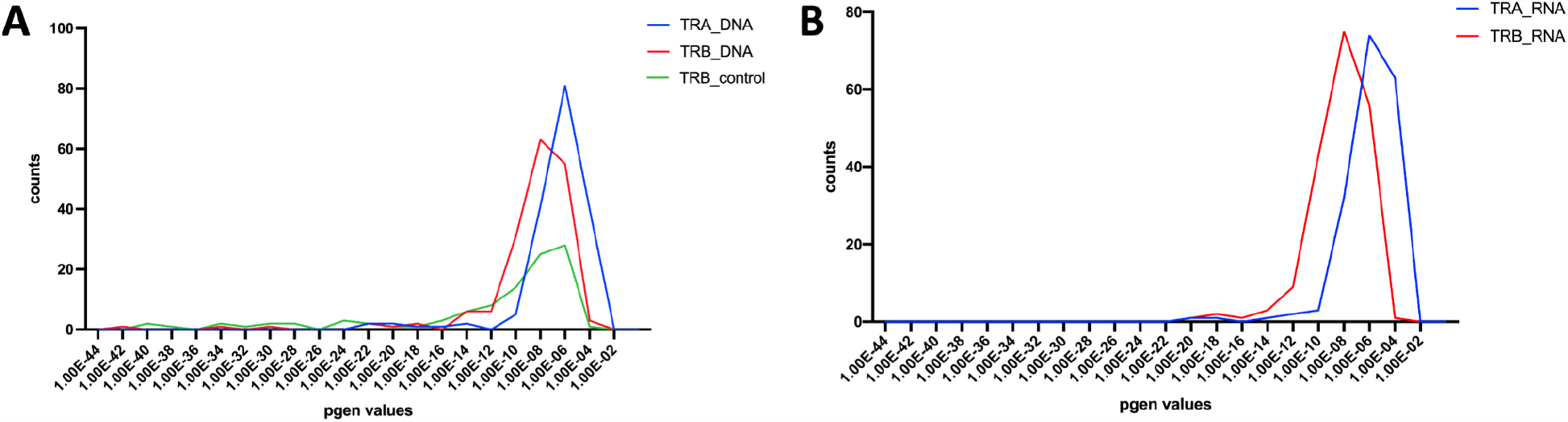
Probability of generation (P_gen_) for TCRα and TCRβ clonotypes. Shared TCRα^aa^ and TCRβ^aa^ clonotypes were identified using the Immunarch pipeline and their P_gen_ was calculated using OLGA algorithm (Fig 1) from DNA and RNA data. **A** - P_gen_ histogram for the shared TCRα^DNA^ and TCRβ^DNA^ clonotypes. As a comparison, the P_gen_ values for the TCRβ^DNA^ clonotypes for the healthy control group are plotted. **B** - analogous histogram as in (A) showing P_gen_ for the shared clonotypes in TCRα^RNA^ and TCRβ^RNA^.

## Discussion

TCLs represent a clinically diverse spectrum of T-cells cancers, varying from aggressive to relatively indolent and from nodal to organ specific (such as the skin). Nevertheless, analysis of TCR rearrangement patterns revealed striking similarities across different TCLs. Because TCR loci are rearranged sequentially and the rearrangement happens only during the thymic development of T-cell precursors, the rearrangement products retained in the genome allow to trace the clonal history of T-cell lymphoma and illuminate the much debated question of the cell of origin.

The current model of TCLs as neoplasms originating from various subpopulations of normal, mature T-cells [3] is not only unsupported, but directly contradicted by our findings. Malignant transformation of a single lymphocyte randomly drawn from a vast pool of 10^6^-10^7^ singular T-cell clonotypes should yield lymphomas which are monoclonal (all malignant cells share the same clonotype), unique (low probability of shared clonotypes between tumors from unrelated patients) and unconnected (random frequency distribution of clonotypes). We show here that TCLs are mostly oligoclonal, have a high degree of interindividual clonotype sharing across different disease entities, and represent connected systems of clonotypes the frequency of which follows the Zipf-Mandelbrot power law.

We propose that considering T-cell lymphatic precursor as a cell of origin allows not only for a more accurate interpretation of our findings but also accounts for previous observations which could not be explained by a mature T-cell model.

Clonotypic diversity arises if a transformed lymphoid precursor passes through different stages of TCR rearrangements resulting in a clonotypically heterogeneous pool of early cancer cells, mimicking the differentiation of normal T-cells. The model schematically shown in **Figure 4 A**, assumes that precursor cell for mature TCLs progress through the sequential rearrangements, analogously to normal thymocytes and in a stark contrast to immature T-cell lymphomas and leukemias (e.g. T-ALL) which become ‘arrested’’ at a given stage of development. Thus, transformation before TCRγ rearrangement would result in oligoclonality for all TCR rearrangements whereas transformation after TCRγ rearrangement would render cancer cells monoclonal for TCRγ but oligoclonal for TCRβ and TCRα. Transformation after the TCRβ rearrangement stage would render the malignant cells monoclonal (single TCRβ^RNA^ clonotype and 1-2 TCRβ^DNA^ clonotypes) but oligoclonal for TCRα. Across different TCLs, ALCL showed a fingerprint of transformation at the earliest stage (before or et the TCR γ rearrangement) and ATCLL and PTCL seemed to derive from a cell in a later stage of development (TCR β rearrangement or later). However, significant overlap in the patterns of clonality existed within each disease indicating that the stage of the transformation does not fully define the phenotype of the disease.

Existence of more than one malignant clonotype has previously been shown in TCLs, but this finding has largely been ignored as artifacts caused by contamination of the sample with normal reactive T-cells. We consider those explanations unlikely. First, in our analysis we include only the most abundant clonotypes of cumulative frequency equal to the proportion of tumor cells in the sample (TCF). Second, across a large number of samples we demonstrated the power-law relationship between the frequency and the rank of the clonotype. Power-law distribution characterizes complex, evolving, out-of-equilibrium, multiplicative systems composed of interconnected components, such as cities in a country, words in the language, repertoire of protein domains, or migrating human populations [43,45,51,52]. It has been shown that growing malignant tumors demonstrate power law distribution between tumor subclones [44]. The distribution of clonotypes in TCLs followed power-law distribution as well, which is only understandable if TCLs are considered as evolving systems of interacting and expanding T-cell clones identified by their clonotypic signature.

We considered alternative explanations of our findings as it could be argued that clonotypic oligoclonality might also be a result of secondary TCR rearrangements in the already established tumor. This is however unlikely as the TCR rearrangement machinery is irreversibly inactivated after completion of TCRα rearrangement and the essential enzymes RAG1 and RAG2 are not expressed in TCLs [53]. Moreover, additional supportive evidence for the lymphoid precursor model for TCLs comes from the pattern of V-J usage and clonotype sharing. TCR rearrangements in human thymocytes are characterized by biased V and J segment usage reflecting the probability of recombination (P_gen_) producing shared (public) clonotypes [54,55]. A very similar situation is seen in TCLs. The V segments preferentially recombined in TCLs (*TRBV20-1, TRBV4-1, TRBV5-1, TRAV14 or TRAV35, TRAV41*) were also the most frequently used segments in normal human thymocytes and the shared clonotypes had comparable, high P_gen_ values (>10^−7^ for TCR α and >10^−9^ for TCRβ) [54,55]. Differences in combinatorial probability is a probable explanation for a higher Jaccard index (degree of clonotype sharing) for TCRα than TCRβ.

It has been postulated that V-J bias in some TCLs is a result of selection pressure from common antigens [56–60]. For example, *TRBV20-1* has the highest frequency across all TCLs and is known to be involved in response to Epstein-Barr or cytomegalovirus [46,61]. However, none of the known CDR3 regions recognizing viral antigen was found among the malignant clonotypes. We postulate that the frequent finding of *TRBV20-1* in TCLs is not due to its antigenic selection but simply its ubiquity. *TRBV20-1* is the prominent component of the TCR β clonotypes of normal blood [62,63], tumor-infiltrating T-cells [64,65], or autoimmune reactions [66–69]. *TRBV20-1* is the most frequently rearranged segment in DP thymocytes [55] and is frequently found in T-cell acute lymphoblastic leukemia (T-ALL) which directly develops from lymphoid precursors [15].

Although our data strongly indicate the role of the lymphoid precursor as the cell of origin in TCLs, the origin of those cells and their pathways of development cannot be inferred from this study. Compelling experimental data suggest that the precursor T-cell in the thymus might be the cell of origin for ALCL [13,14], but we cannot exclude the possibility that extrathymic sites are involved as well. T-cell maturation and TCR recombination may happen in extrathymic sites such as bone marrow, lymph nodes or the tonsils [70–72]. We have found early MF precursor cells in the bone marrow before the clinical onset of the disease [73] and cases of transmission of T-cell lymphoma by bone marrow transplant from asymptomatic donors who later developed the disease have been observed [74] indicating that bone marrow could be a potential reservoir or progenitor malignant cells.

In summary, we propose that mature TCLs do not originate from the T-cells in the periphery but rather from lymphoid precursors at different stages of their differentiation. Better understanding of the developmental trajectories of the early precursor into clinical lymphoid malignancy would improve diagnosing and staging of TCLs and help to design targeted therapies intervening with the disease at the initial stages.

## Data Availability

The data accession number for all studies have been listed in supplementary table 1.

## Acknowledgements

The authors would like to thank the NIH Data access committee and EGA European Genome-Phenome Archive database for their help with getting access to certain restricted datasets.

This research was funded by the unrestricted grants from the University of Alberta (Canada) and Bispebjerg Hospital (Copenhagen, Denmark), Danish Cancer Society (Kræftens Bekæmpelse; R124-A7592 Rp12350), Canadian Dermatology Foundation (CDF) and University Hospital Foundation (University of Alberta). Additionally Dr. Aishwarya Iyer was funded under the accelerate postdoctoral fellowship under a joint grant from the Mitacs and Sun Pharma Global FZE.

## Competing interests

All authors have completed the ICMJE uniform disclosure form and declare: RG - no support from any organisation for the submitted work; speaker honoraria and advisory board honoraria from the following organisations that might have an interest in the submitted work - Mallinckrodt, Lilly, Sanofi, Abbvie, Sunpharma, Kyowa Kirin; no other relationships or activities that could appear to have influenced the submitted work. Other authors - no competing interests.

## Notes

### Clinical Trial

The trial though collects human genome data does not administer any treatment that requires a clinical trial registration.

### Author Declarations

To access the human genome data from restricted databases such as dbGaP and EGA was provided by Health Research Ethics Board of Alberta Cancer Committee under ethics ID- HREBA.CC-20-0118.

