## Supplementary tables for "Clonotype pattern in T-cell lymphomas map the cell of origin to immature lymphoid precursors"

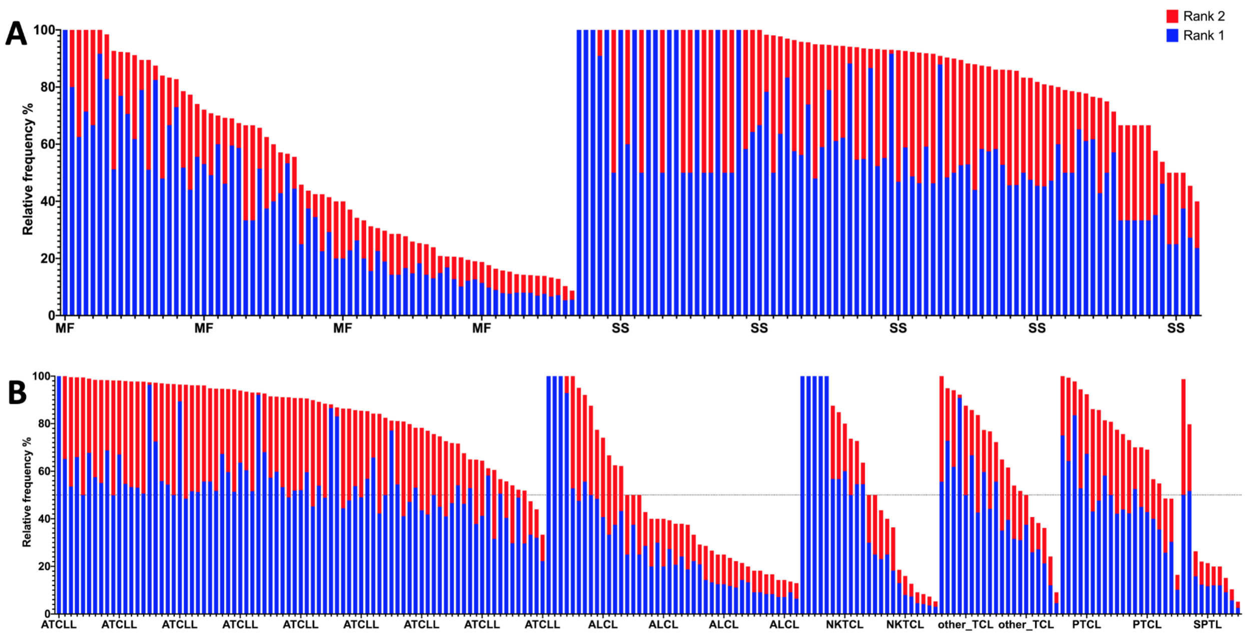


**Supplementary Fig S1. Frequency of TCRγ clonotypes.**
TCRγ clonotype frequency was identified in 378 DNA samples from various subgroups of TCL. The frequency of 2 most abundant clonotypes were plotted as bar graphs. TCRγ clonotype frequency for (A) MF and SS samples (B) other subgroups of TCL. MF-mycosis fungoides; SS- sezary syndrome; ATCLL- adult T-cell lymphoma and leukemia; ALCL- anaplastic large cell lymphoma; NKTL- natural killer T-cell lymphoma; PTCL- peripheral T-cell lymphoma and SPTL- subcutaneous panniculitis-like T-cell lymphoma.

**Table S1**
List of studies and number of samples included in the meta-analysis study.

| Study | Type of T-cell lymphoma | Number of samples | SRA or dbGaP reference |
|---|---|---|---|
| Iyer et al.[1] | CTCL | WES (58), WTS (42) | phs001877.v1.p1 |
| Choi et al.[2] | CTCL | WES (34) | SRP058948 |
| Ungewickell et al.[3] | CTCL | WES (73) | phs000913.v1.p1 |
| McGirt et al.[4] | CTCL | WGS (5) | SRP059214 |
| Almeida et al.[5] | CTCL | WES (40) | phs000994.v1.p1 |
| Li et al.[5,6] | SPTL | WES (10) | SRP078777 |
| Simpson et al.[7] | PTCL | WES (12) | SRP047386 |
| Palomero et al.[8] | PTCL | WES (12), WTS (35) | phs000689.v1.p1 |

| | | | |
|---|---|---|---|
| Wang et al.[9] | CTCL | WES (37), WTS (32) | phs000859.v1.p1 |
| Yoo et al.[10] | ATCL | WES (7), WTS (10) | SRP029591 |
| Jiang et al.[11] | NKTL | WES (25) | SRP057085 |
| Crescenzo et al.[12] | ALCL | WES (47), WTS (20) | SRP044708 |
| Kataoka et al.[13] | ATCLL | WES (81), WTS (60) | EGAS00001001296 |

CTCL (cutaneous T-cell lymphoma), SPTL (subcutaneous panniculitis‑like T‑cell lymphoma), PTCL (Peripheral T Cell Lymphoma), ATCL (angioimmunoblastic T cell lymphoma), NKTL (natural killer/T-cell lymphoma), ALCL ( Anaplastic Large Cell Lymphoma), ATCLL (adult T cell leukemia/lymphoma), WES (whole exome sequencing), WTS (whole transcriptome sequencing), WGS (whole genome sequencing).

**Table S2**
List of samples grouped as other TCLs

| Sample | Type of TCL | Sample Type |
|---|---|---|
| Pt13 | T-cell large granular lymphocytic leukemia | DNA |
| Pt14 | T-cell prolymphocytic leukemia | DNA |
| Pt6 | Hepatosplenic T-cell lymphoma | DNA |
| Pt4 | Anaplastic large cell lymphoma unspecified | DNA |
| NU1T | Cutaneous T-cell lymphoma | DNA |
| NU3T | Non MF or SS Cutaneous T-cell lymphoma | DNA |
| NU6T | Non MF or SS Cutaneous T-cell lymphoma | DNA |
| NU12T | Non MF or SS Cutaneous T-cell lymphoma | DNA |
| NU13T | Non MF or SS Cutaneous T-cell lymphoma | DNA |
| NU14T | Non MF or SS Cutaneous T-cell lymphoma | DNA |
| NU16T | Non MF or SS Cutaneous T-cell lymphoma | DNA |
| NU17T | Non MF or SS Cutaneous T-cell lymphoma | DNA |
| NU21T | Non MF or SS Cutaneous T-cell lymphoma | DNA |
| PAT1 | Angioimmunoblastic T-cell lymphoma | RNA |

| | | |
|---|---|---|
| PAT2 | Angioimmunoblastic T-cell lymphoma | RNA |
| PAT3 | Angioimmunoblastic T-cell lymphoma | RNA |
| PAT4 | Angioimmunoblastic T-cell lymphoma | RNA |
| PAT5 | Angioimmunoblastic T-cell lymphoma | RNA |
| PAT6 | Angioimmunoblastic T-cell lymphoma | RNA |
| PAT7 | Angioimmunoblastic T-cell lymphoma | RNA |
| PAT8 | Angioimmunoblastic T-cell lymphoma | RNA |
| PAT9 | Angioimmunoblastic T-cell lymphoma | RNA |
| PAT13 | Angioimmunoblastic T-cell lymphoma | RNA |

**Table S3**
Tumor cell fraction and frequency of the most abundant TCRβ clonotype.

| Sample | Sample Type | TCF | $TCR\beta^{DNA}$ clonotype |
|---|---|---|---|
| MF19_1 | MF | 98.573 | 5.555556 |
| MF45.1 | MF | 96.368 | 14.47368 |
| MF19_3 | MF | 96.316 | 15.90909 |
| MF8.1 | MF | 92.583 | 15.87302 |
| MF27.1 | MF | 92.312 | 25.18519 |
| MF10.1 | MF | 92.182 | 25.71429 |
| MF29_2 | MF | 92.178 | 17.3913 |
| MF15.1 | MF | 91.966 | 9.009009 |
| MF12.1 | MF | 91.397 | 31.46067 |
| MF2.1 | MF | 91.309 | 25 |
| MF4_4 | MF | 91.177 | 50.20747 |
| MF9.1 | MF | 90.173 | 17.24138 |
| MF20.1 | MF | 90.167 | 13.39286 |
| MF42Der | MF | 89.93 | 28.04878 |
| MF17Der | MF | 89.47 | 26.08696 |

| | | | |
|---|---|---|---|
| MF29_1 | MF | 89.26 | 10.37736 |
| MF4_1 | MF | 87.97 | 33.33333 |
| MF25.1 | MF | 85.27 | 9.243697 |
| MF28Der | MF | 83.17 | 4.065041 |
| MF11.1 | MF | 82.29 | 17.91045 |
| MF31.1 | MF | 81.08 | 10.34483 |
| MF4_7 | MF | 80.88 | 61.76471 |
| MF4_5 | MF | 80.36 | 75.24272 |
| MF32_1 | MF | 80.14 | 64.13793 |
| p3 | MF | 75.73 | 50 |
| MF22Der | MF | 73.7 | 44.44444 |
| MF4_3 | MF | 73.37 | 30.46358 |
| MF4_6 | MF | 72.37 | 22.58065 |
| MF7_1 | MF | 71.28 | 9.375 |
| 7 | MF | 68.6 | 20 |
| MF19_2 | MF | 67.27 | 9.756098 |
| MF4_2 | MF | 66.79 | 19.7479 |
| MF32.1 | MF | 65.27 | 10 |
| MF30.1 | MF | 62.89 | 14.44444 |
| MF7_2 | MF | 62.03 | 8.571429 |
| MF18Der | MF | 60.52 | 28.23129 |
| MF39_1 | MF | 56.34 | 30 |
| MF5_1 | MF | 55.99 | 10 |
| MF11_1 | MF | 55.58 | 10 |
| MF11_2 | MF | 55.45 | 10.44776 |
| MF38_2 | MF | 55.18 | 23.3871 |
| MF34_1 | MF | 54.85 | 28.91566 |
| MF26.1 | MF | 50.49 | 14.60674 |
| NU7T | MF | 50.43 | 23.91304 |
| 5 | MF | 49.69 | 15.25424 |
| NU20T | MF | 49.45 | 10.20408 |

| | | | |
|---|---|---|---|
| 9 | MF | 48.96 | 7.142857 |
| MF37.1 | MF | 48.53 | 14.70588 |
| MF38_1 | MF | 48.19 | 8.955224 |
| MF16.1 | MF | 46.45 | 7.307172 |
| MF41Der | MF | 44.73 | 12.19512 |
| 16 | MF | 44.08 | 5.882353 |
| MF23Der | MF | 42.95 | 12.96296 |
| NU19T | MF | 39.3 | 25.89286 |
| MF5_2 | MF | 35.67 | 6.818182 |
| MF44.1 | MF | 35.59 | 17.56757 |
| p4 | MF | 33.17 | 100 |
| MF34_2 | MF | 32.37 | 16.30435 |
| MF35.1 | MF | 31.22 | 17.3913 |
| p1 | MF | 29.72 | 91.66667 |
| MF34.1 | MF | 29.57 | 14.60674 |
| MF36.1 | MF | 28.24 | 4.411765 |
| MF40_2 | MF | 25.95 | 13.18681 |
| NU4T | MF | 25.56 | 29.56522 |
| p2 | MF | 24.43 | 75 |
| MF23_1 | MF | 23.88 | 5.445545 |
| 8 | MF | 23.38 | 55.88235 |
| MF40_1 | MF | 21.94 | 11.34021 |
| NU2T | MF | 21.46 | 6.451613 |
| MF43.1 | MF | 21.14 | 10.20408 |
| p5 | MF | 12.86 | 100 |
| NU9T | MF | 11.13 | 5 |
| 52 | MF | 9.87 | 23.91304 |
| NU18T | MF | 0.93 | 5.042017 |
| NU11T | MF | 0.39 | 3.448276 |
| WES-CTCL11-tumor | SS | 99 | 7.594937 |
| WES-CTCL12-tumor | SS | 99 | 6.578947 |

| | | | |
|---|---|---|---|
| WES-CTCL23-tumor | SS | 99 | 11.68831 |
| WES-CTCL6-tumor | SS | 98 | 22.72727 |
| WES-CTCL9-tumor | SS | 98 | 15 |
| WES-CTCL13-tumor | SS | 98 | 6.976744 |
| 029-CD4-01T-1 | SS | 97.347 | 8.695652 |
| 009-CD4-01T-1 | SS | 97.212 | 9.090909 |
| WES-CTCL39-tumor | SS | 96 | 5.952381 |
| 021-CD4-01T-D-1 | SS | 95.685 | 19.44444 |
| 4 | SS | 95.354 | 50.76923 |
| 032-CD4-01T-1 | SS | 95.287 | 37.5 |
| WES-CTCL2-tumor | SS | 95 | 10.66667 |
| WES-CTCL21-tumor | SS | 95 | 12.22222 |
| WES-CTCL22-tumor | SS | 95 | 7.5 |
| 036-CD4-01T-1 | SS | 94.027 | 6.060606 |
| WES-CTCL3-tumor | SS | 94 | 13.11475 |
| WES-CTCL4-tumor | SS | 94 | 20 |
| WES-CTCL8-tumor | SS | 94 | 14.47368 |
| WES-CTCL20-tumor | SS | 93 | 5.319149 |
| WES-CTCL29-tumor | SS | 93 | 7.017544 |
| WES-CTCL36-tumor | SS | 93 | 16.07143 |
| WES-CTCL40-tumor | SS | 93 | 11.66667 |
| Pt7 | SS | 92.77 | 56.66667 |
| 017-CD4-01T-D-1 | SS | 92.534 | 7.692308 |
| WES-CTCL1-tumor | SS | 92 | 12.98701 |
| 018-CD4-01T-D-1 | SS | 91.798 | 31.03448 |
| 020-CD4-01T-D-1 | SS | 91.513 | 3.846154 |
| 005-CD4-01T-1 | SS | 90.806 | 9.52381 |
| 022-CD4-01T-D-1 | SS | 90.778 | 8.333333 |
| WES-CTCL19-tumor | SS | 90 | 9.756098 |
| 030-CD4-01T-1 | SS | 89.16 | 29.54545 |
| 15 | SS | 87.43 | 28 |

| | | | |
|---|---|---|---|
| 014-CD4-01T-1 | SS | 86.83 | 46.15385 |
| WES-CTCL35-tumor | SS | 85 | 14.58333 |
| 002-CD4-01T-1 | SS | 84.17 | 8.695652 |
| WES-CTCL30-tumor | SS | 81 | 16.07143 |
| 034-CD4-01T-1 | SS | 74.56 | 6.451613 |
| 012-CD4-01T-1 | SS | 72.58 | 26.66667 |
| 033-CD4-01T-1 | SS | 72.5 | 8.695652 |
| WES-CTCL38-tumor | SS | 72 | 18.75 |
| 031-CD4-01T-1 | SS | 71.57 | 4.761905 |
| 016-CD4-01T-1 | SS | 71.26 | 3.703704 |
| 1 | SS | 71.17 | 21.875 |
| NU8T | SS | 68.09 | 18.51852 |
| WES-CTCL7-tumor | SS | 63 | 11.00917 |
| 003-CD4-01T-1 | SS | 62.57 | 5.555556 |
| 015-CD4-01T-1 | SS | 62.11 | 3.448276 |
| 011-CD4-01T-1 | SS | 59.23 | 6.666667 |
| NU15T | SS | 56.23 | 9.52381 |
| 024-CD4-01T-D-1 | SS | 52.92 | 25 |
| 54 | SS | 52.88 | 16.66667 |
| 23B | SS | 51.95 | 2.439024 |
| 027-CD4-01T-1 | SS | 51.04 | 7.692308 |
| 11 | SS | 50.52 | 35.29412 |
| 026-CD4-01T-D-1 | SS | 47.23 | 6.666667 |
| WES-CTCL5-tumor | SS | 47 | 8.695652 |
| 5B | SS | 46.41 | 6.493506 |
| WES-CTCL15-tumor | SS | 44 | 4.83871 |
| 025-CD4-01T-D-1 | SS | 43.09 | 9.090909 |
| WES-CTCL31-tumor | SS | 41 | 9.876543 |
| WES-CTCL33-tumor | SS | 41 | 10.43478 |
| 18B | SS | 39.22 | 3.703704 |
| 17B | SS | 38.57 | 4.83871 |

| | | | |
|---|---|---|---|
| 001-CD4-01T-1 | SS | 37.01 | 10.52632 |
| 21B | SS | 36.96 | 4.316547 |
| 008-CD4-01T-1 | SS | 35.21 | 7.142857 |
| 4B | SS | 34.69 | 11.11111 |
| 22B | SS | 34.26 | 10 |
| WES-CTCL16-tumor | SS | 34 | 6.25 |
| 3B | SS | 33.85 | 14.77273 |
| 16B | SS | 32.09 | 11.90476 |
| 8B | SS | 31.68 | 5 |
| 028-CD4-01T-1 | SS | 29.84 | 3.333333 |
| 023-CD4-01T-D-1 | SS | 29.19 | 9.090909 |
| 11B | SS | 28.56 | 7.692308 |
| 14B | SS | 26.62 | 5.747126 |
| 19B | SS | 26.61 | 6.666667 |
| 038-CD4-01T-1 | SS | 25.84 | 5.555556 |
| 2B | SS | 25.4 | 4.210526 |
| 010-CD4-01T-1 | SS | 25.21 | 4.545455 |
| 007-CD4-01T-1 | SS | 24.91 | 8 |
| 6B | SS | 24.59 | 23.07692 |
| 019-CD4-01T-D-1 | SS | 23.67 | 19.23077 |
| 9B | SS | 23.22 | 4.761905 |
| 1B | SS | 22.53 | 15.71429 |
| 7B | SS | 19.94 | 7.894737 |
| 15B | SS | 14.63 | 30.53435 |
| 10B | SS | 11.04 | 4.938272 |
| 037-CD4-01T-1 | SS | 6.27 | 4.347826 |
| WES-CTCL10-tumor | SS | 4 | 23.91304 |
| WES-CTCL17-tumor | SS | 4 | 6.666667 |
| WES-CTCL32-tumor | SS | 2 | 9.090909 |
| 12B | SS | 0.45 | 11.25 |
| 20B | SS | 0.4 | 23.72881 |

| | | | |
|---|---|---|---|
| 13B | SS | 0.32 | 7.936508 |
| WES-CTCL18-tumor | SS | 0 | 13.7931 |
| WES-CTCL26-tumor | SS | 0 | 16.66667 |
| ATL070_T | ATCLL | 97.549 | 40.2985075 |
| ATL002_T | ATCLL | 96.172 | 26.1904762 |
| ATL016_T | ATCLL | 95.26 | 38.0952381 |
| ATL023_T | ATCLL | 94.834 | 9.09090909 |
| ATL089_T | ATCLL | 93.348 | 38.5620915 |
| ATL088_T | ATCLL | 90.138 | 27.4509804 |
| ATL069_T | ATCLL | 89.99 | 56.7901235 |
| ATL015_T | ATCLL | 88.65 | 40.1709402 |
| ATL058_T | ATCLL | 88.38 | 46.25 |
| ATL052_T | ATCLL | 88.09 | 44.6043165 |
| ATL026_T | ATCLL | 86.84 | 37.1428571 |
| ATL040_T | ATCLL | 86.13 | 42.5925926 |
| ATL071_T | ATCLL | 84.46 | 10.7142857 |
| ATL030_T | ATCLL | 83.79 | 42.7184466 |
| ATL021_T | ATCLL | 83.47 | 52.9850746 |
| ATL046_T | ATCLL | 83.47 | 24.2424242 |
| ATL022_T | ATCLL | 83.12 | 14.2857143 |
| ATL033_T | ATCLL | 79.36 | 65.0406504 |
| ATL014_T | ATCLL | 78.42 | 35.5769231 |
| ATL008_T | ATCLL | 78.32 | 58.974359 |
| ATL065_T | ATCLL | 78.23 | 50 |
| ATL018_T | ATCLL | 75.31 | 36.6120219 |
| ATL034_T | ATCLL | 74.21 | 25 |
| ATL090_T | ATCLL | 72.98 | 39.6825397 |
| ATL038_T | ATCLL | 72.62 | 11.3636364 |
| ATL035_T | ATCLL | 68.2 | 44.4444444 |
| ATL081_T | ATCLL | 66.53 | 11.1111111 |
| ATL027_T | ATCLL | 66.16 | 16.3265306 |

| | | | |
|---|---|---|---|
| ATL050_T | ATCLL | 63.87 | 33.3333333 |
| ATL055_T | ATCLL | 63.86 | 13.559322 |
| ATL011_T | ATCLL | 62.73 | 38.75 |
| ATL020_T | ATCLL | 61.51 | 34.0206186 |
| ATL076_T | ATCLL | 60.69 | 6.25 |
| ATL074_T | ATCLL | 59.67 | 20 |
| ATL032_T | ATCLL | 59.03 | 18.6813187 |
| ATL039_T | ATCLL | 58.33 | 6.81818182 |
| ATL037_T | ATCLL | 57.27 | 43.75 |
| ATL029_T | ATCLL | 56.8 | 30.3797468 |
| ATL075_T | ATCLL | 56.34 | 67.0588235 |
| ATL007_T | ATCLL | 56.07 | 25 |
| ATL048_T | ATCLL | 54.94 | 25 |
| ATL043_T | ATCLL | 52.04 | 21.4285714 |
| ATL012_T | ATCLL | 51.49 | 21.0526316 |
| ATL044_T | ATCLL | 51.01 | 33.3333333 |
| ATL028_T | ATCLL | 50.82 | 18.1818182 |
| ATL059_T | ATCLL | 49.24 | 14.1509434 |
| ATL079_T | ATCLL | 45.8 | 22.4489796 |
| ATL047_T | ATCLL | 44.9 | 17.6470588 |
| ATL057_T | ATCLL | 43.05 | 39.0243902 |
| ATL062_T | ATCLL | 41.85 | 31.5789474 |
| ATL085_T | ATCLL | 40.75 | 15.7894737 |
| ATL031_T | ATCLL | 40.66 | 7.84313725 |
| ATL001_T | ATCLL | 39.21 | 21.2389381 |
| ATL005_T | ATCLL | 34.42 | 23.4375 |
| ATL036_T | ATCLL | 31.48 | 21.6666667 |
| ATL013_T | ATCLL | 29.35 | 12.987013 |
| ATL049_T | ATCLL | 29.32 | 23.75 |
| ATL045_T | ATCLL | 28.72 | 47.1830986 |
| ATL067_T | ATCLL | 26.3 | 10 |

| | | | |
|---|---|---|---|
| ATL072_T | ATCLL | 25.17 | 5.26315789 |
| ATL051_T | ATCLL | 24.95 | 44.5945946 |
| ATL041_T | ATCLL | 24.94 | 46.6666667 |
| ATL077_T | ATCLL | 23.77 | 42.3529412 |
| ATL004_T | ATCLL | 20.95 | 26.5822785 |
| ATL025_T | ATCLL | 20.8 | 38.4615385 |
| ATL063_T | ATCLL | 19.52 | 12.3287671 |
| ATL056_T | ATCLL | 18.19 | 16.6666667 |
| ATL073_T | ATCLL | 15.35 | 17.3913043 |
| ATL017_T | ATCLL | 14.81 | 12 |
| ATL068_T | ATCLL | 11.16 | 6.97674419 |
| ATL010_T | ATCLL | 8.82 | 40.1515152 |
| ATL084_T | ATCLL | 7.5 | 20.3703704 |
| ATL078_T | ATCLL | 7.09 | 29.2682927 |
| ATL083_T | ATCLL | 6.83 | 62.8205128 |
| ATL009_T | ATCLL | 6.28 | 31.372549 |
| ATL054_T | ATCLL | 4.62 | 27.1317829 |
| ATL024_T | ATCLL | 3.73 | 46.969697 |
| ATL019_T | ATCLL | 3.34 | 51.8796992 |
| ATL053_T | ATCLL | 0.82 | 42.3076923 |
| ATL064_T | ATCLL | 0.44 | 7.5 |
| ATL060_T | ATCLL | 0.02 | 26.984127 |
| GPS60 | ALCL | 90.07 | 12.5 |
| GPS63 | ALCL | 78.56 | 31.8181818 |
| GPS67 | ALCL | 60.97 | 10 |
| GPS71 | ALCL | 60.84 | 4.76190476 |
| GPS73 | ALCL | 60.8 | 9.09090909 |
| GPS41 | ALCL | 60.59 | 8.69565217 |
| GPS59 | ALCL | 60.08 | 28.9473684 |
| GPS77 | ALCL | 59.16 | 7.14285714 |
| GPS79 | ALCL | 59 | 8.69565217 |

| | | | |
|---|---|---|---|
| GPS56 | ALCL | 58.93 | 5 |
| GPS94 | ALCL | 58.91 | 14.2857143 |
| GPS72 | ALCL | 58.8 | 3.22580645 |
| GPS90 | ALCL | 58.76 | 13.3333333 |
| GPS64 | ALCL | 58.55 | 3.33333333 |
| GPS76 | ALCL | 58.51 | 8 |
| GPS33 | ALCL | 57.39 | 11.7647059 |
| GPS74 | ALCL | 57.3 | 15 |
| GPS61 | ALCL | 56.8 | 15.3846154 |
| GPS36 | ALCL | 56.61 | 5.26315789 |
| GPS37 | ALCL | 56.27 | 5.88235294 |
| GPS69 | ALCL | 56.01 | 12 |
| GPS39 | ALCL | 55.85 | 16.6666667 |
| GPS32 | ALCL | 55.81 | 6.66666667 |
| GPS40 | ALCL | 55.74 | 9.09090909 |
| GPS27 | ALCL | 55.71 | 11.1111111 |
| GPS38 | ALCL | 55.48 | 7.40740741 |
| GPS78 | ALCL | 55.47 | 7.69230769 |
| GPS21 | ALCL | 55.19 | 8.33333333 |
| GPS25 | ALCL | 55.19 | 9.52380952 |
| GPS89 | ALCL | 55.14 | 10 |
| GPS93 | ALCL | 55.14 | 14.2857143 |
| GPS66 | ALCL | 54.88 | 17.6470588 |
| GPS65 | ALCL | 54.74 | 3.44827586 |
| GPS68 | ALCL | 54.47 | 12.5 |
| GPS70 | ALCL | 54.42 | 6.4516129 |
| GPS75 | ALCL | 54.18 | 5 |
| GPS22 | ALCL | 53.75 | 8.33333333 |
| GPS58 | ALCL | 53.64 | 14.2857143 |
| GPS28 | ALCL | 53.02 | 8.69565217 |
| GPS62 | ALCL | 52.95 | 11.1111111 |

| | | | |
|---|---|---|---|
| GPS57 | ALCL | 52.72 | 15.3846154 |
| GPS46 | ALCL | 52.13 | 13.6363636 |
| GPS42 | ALCL | 51.54 | 30 |
| GI11 | ALCL | 43.28 | 26.3157895 |
| GPS24 | ALCL | 14.84 | 5.40540541 |
| GPS23 | ALCL | 11.1 | 9.67741935 |
| GI14 | ALCL | 0 | 8.69565217 |
| L08A | NKTCL | 83.06 | 40 |
| L20A | NKTCL | 78.04 | 4.16666667 |
| L22A | NKTCL | 70.81 | 26.0869565 |
| L03A | NKTCL | 68.4 | 40 |
| L15A | NKTCL | 61.77 | 5.69105691 |
| L02A | NKTCL | 61.75 | 20 |
| L12A | NKTCL | 51.82 | 15.3846154 |
| L24A | NKTCL | 50.27 | 4.87804878 |
| L14A | NKTCL | 47.74 | 11.1111111 |
| L01A | NKTCL | 45.3 | 26.6666667 |
| L18A | NKTCL | 39.64 | 8.82352941 |
| L13A | NKTCL | 39.34 | 16.6666667 |
| L07A | NKTCL | 37.5 | 11.1111111 |
| L21A | NKTCL | 36.09 | 7.57575758 |
| L23A | NKTCL | 34.26 | 6.06060606 |
| L09A | NKTCL | 32.66 | 14.2857143 |
| L05A | NKTCL | 31.29 | 10.5263158 |
| L17A | NKTCL | 19.56 | 5.35714286 |
| L16A | NKTCL | 14.41 | 5.55555556 |
| L06A | NKTCL | 10.67 | 15 |
| L25A | NKTCL | 10.59 | 7.31707317 |
| L10A | NKTCL | 9.15 | 33.3333333 |
| L19A | NKTCL | 8.62 | 6 |
| L04A | NKTCL | 6.22 | 30 |

| | | | |
|---|---|---|---|
| L11A | NKTCL | 0.39 | 14.2857143 |
| Pt14 | other_TCL | 97.714 | 43.9393939 |
| Pt6 | other_TCL | 97.703 | 15 |
| PAT3 | other_TCL | 81.15 | 5.66037736 |
| NU14T | other_TCL | 79.05 | 57.2033898 |
| NU13T | other_TCL | 76.59 | 18.8405797 |
| PAT10 | other_TCL | 60.29 | 38.9830508 |
| NU17T | other_TCL | 32.37 | 11.7117117 |
| PAT11 | other_TCL | 28.84 | 5.88235294 |
| Pt13 | other_TCL | 26.64 | 64.516129 |
| PAT5 | other_TCL | 26.22 | 6.81818182 |
| NU1T | other_TCL | 25.41 | 15.2777778 |
| PAT1 | other_TCL | 24.8 | 29.2682927 |
| Pt4 | other_TCL | 22.9 | 71.1864407 |
| NU3T | other_TCL | 16.55 | 2.43902439 |
| NU21T | other_TCL | 16.53 | 4.12371134 |
| NU16T | other_TCL | 10.8 | 3.57142857 |
| PAT12 | other_TCL | 0.42 | 22.5806452 |
| PAT4 | other_TCL | 0.33 | 14.4927536 |
| NU12T | other_TCL | 0.16 | 5.47945205 |
| NU6T | other_TCL | 0 | 3.7037037 |
| Pt11 | PTCL | 99.3384 | 56.25 |
| 33T | PTCL | 95.675 | 51.3888889 |
| Pt19_1 | PTCL | 94.996 | 53.3333333 |
| Pt10 | PTCL | 94.492 | 48.9361702 |
| 29T | PTCL | 93.953 | 11.5384615 |
| 26T | PTCL | 85.56 | 5.97014925 |
| Pt21 | PTCL | 73.35 | 7.46268657 |
| Pt19_2 | PTCL | 68.02 | 80 |
| 2T | PTCL | 64.12 | 12.5 |
| Pt17 | PTCL | 60.28 | 4.76190476 |

| | | | |
|---|---|---|---|
| 24T | PTCL | 48.71 | 9.25925926 |
| Pt12 | PTCL | 39.27 | 40.3846154 |
| 1T | PTCL | 36.17 | 12.5 |
| 31T | PTCL | 34.47 | 31.7460317 |
| 28T | PTCL | 31.42 | 26.3157895 |
| 5T | PTCL | 29.16 | 9.25925926 |
| 11T | PTCL | 27.6 | 7.40740741 |
| 4T | PTCL | 22.57 | 43.4782609 |
| 35T | PTCL | 22.33 | 45.4545455 |
| Pt26 | PTCL | 1.48 | 43.0769231 |
| SP3 | SPTCL | 66.74 | 94.0397351 |
| SP13 | SPTCL | 40.64 | 7.14285714 |
| SP12 | SPTCL | 36.76 | 25 |
| SP15 | SPTCL | 30.5 | 7.69230769 |
| SP10 | SPTCL | 23.78 | 18.1818182 |
| SP4 | SPTCL | 22.31 | 8.33333333 |
| SP14 | SPTCL | 19.93 | 10 |
| SP8 | SPTCL | 10.01 | 19.4444444 |
| SP7 | SPTCL | 9.365 | 8 |
| SP1 | SPTCL | 1.74 | 33.3333333 |

**Table S4**
$P_{gen}$ values for the $TCR\beta^{aa}$ clonotypes that were shared between TCLs.

| Predictive CDR3 amino acid sequence | *TRBV* genes | Number of shared samples | $P_{gen}$ value |
|---|---|---|---|
| CASSYSNQPQHF | TRBV19 | 4 | 7.12E-07 |
| CASSSGANVLTF | TRBV5-1 | 2 | 2.64E-07 |
| CSARTGGYGYTF | TRBV20-1 | 2 | 1.67E-07 |
| CASSYSGGDEQFF | TRBV6-5 | 2 | 7.94E-08 |
| CASSATGTNYGYTF | TRBV6-4 | 3 | 4.00E-08 |
| CARYF | TRBV25-1 | 10 | 2.94E-08 |

| | | | |
|---|---|---|---|
| CASSF | TRBV12-1 | 16 | 1.49E-08 |
| CASRIYF | TRBV12-2 | 14 | 1.39E-08 |
| CASGRDRHTEAFF | TRBV10-2 | 2 | 6.24E-09 |
| CATSRGGHQETQYF | TRBV24-1 | 2 | 5.91E-09 |
| CASSKVTGGTDTQYF | TRBV10-2 | 3 | 3.91E-09 |
| CRGGF | TRBV3-1 | 2 | 3.60E-09 |
| CSAVAGFSYEQYF | TRBV20-1 | 6 | 2.30E-09 |
| CASSEQESSPLHF | TRBV6-4 | 3 | 1.30E-09 |
| CASSPDETGSYEQYF | TRBV7-6 | 2 | 7.87E-10 |
| CASSYKGQGENSPLHF | TRBV6-5 | 6 | 5.20E-10 |
| CASSFSVGPTYEQYF | TRBV14 | 2 | 4.89E-10 |
| CASSSLKGDSYNEQFF | TRBV5-4 | 5 | 4.74E-10 |
| CASSSGTGHVNQPQHF | TRBV6-5 | 2 | 4.66E-10 |
| CIQQFF | TRBV1 | 3 | 4.25E-10 |
| CASSYSTGVALHF | TRBV6-6 | 2 | 3.91E-10 |
| CVIKYF | TRBV10-3 | 12 | 2.24E-10 |
| CSFHF | TRBV20-1 | 2 | 1.13E-10 |
| CAGLGGRDQETQYF | TRBV12-5 | 2 | 7.54E-11 |
| CASSRPWGGFDEQYF | TRBV18 | 3 | 4.90E-11 |
| CGHQFF | TRBV10-3 | 3 | 2.42E-11 |
| LYFF | TRBV6-7 | 8 | 1.92E-11 |
| CASNRDGRAPLHF | TRBV6-5 | 3 | 1.87E-11 |
| CVCLLF | TRBV7-4 | 2 | 1.12E-11 |
| CARRKSSFF | TRBV28 | 54 | 1.01E-11 |
| CASSRPCGGFDEQYF | TRBV18 | 2 | 7.44E-12 |
| WSCLF | TRBV22-1 | 26 | 6.64E-12 |
| CASSIVGVTSYSNQPQHF | TRBV19 | 10 | 4.98E-12 |
| LFHF | TRBV27 | 9 | 3.82E-12 |
| CARHHRDVF | TRBV18 | 4 | 2.28E-12 |
| CASSGMEGQGALYGYTF | TRBV28 | 2 | 2.24E-12 |
| CASRQKWTGAGSPLHF | TRBV6-1 | 3 | 3.10E-13 |

| | | | |
|---|---|---|---|
| CAKQLKLLF | TRBV5-2 | 3 | 1.22E-13 |
| CFRATIFF | TRBV16 | 5 | 8.42E-14 |
| CASSTYRGAAPPQYF | TRBV28 | 4 | 5.33E-14 |
| FIFIHF | TRBV22-1 | 23 | 1.22E-14 |
| CGEQRGTDSFF | TRBV12-3 | 4 | 4.15E-16 |
| CFCLGNVVLTF | TRBV7-9 | 4 | 1.20E-16 |
| SASITPFDF | TRBV13 | 3 | 8.98E-18 |
| LLLLLLLLLFF | TRBV16 | 7 | 4.80E-18 |
| LPVLGQHVATQHF | TRBV5-6 | 3 | 3.29E-20 |
| CASIEDLITVGHLYLF | TRBV3-1 | 2 | 2.40E-22 |
| CDISQHYDHGFQYF | TRBV10-3 | 29 | 8.58E-23 |
| CAHLLLTSRIHDGGFF | TRBV23-1 | 3 | 2.36E-23 |
| CREAVCGCSASLSF | TRBV1 | 4 | 1.18E-23 |
| WHPSHLLFFFFVF | TRBV6-7 | 14 | 4.89E-24 |
| CLARDVSDVRNYRIYF | TRBV6-9 | 10 | 4.49E-25 |
| WEGLLSPFHLCRFHF | TRBV14 | 3 | 5.47E-26 |
| CGSSFQAEQPITARCEQFF | TRBV4-1 | 8 | 2.40E-26 |
| CVQQHVNVYSTTFFF | TRBV21-1 | 11 | 1.90E-26 |
| CLDFVKIKVVIWLIF | TRBV3-1 | 5 | 1.53E-26 |
| CHAETSQGRFYGIIHF | TRBV22-1 | 2 | 8.07E-27 |
| CAVHHTALPGAQGPTSAF | TRBV18 | 3 | 6.21E-27 |
| CAPHLSTLHHVPHQPLF | TRBV4-1 | 3 | 4.48E-27 |
| CASERRYSASARRLCISRATF | TRBV4-3 | 3 | 5.48E-30 |
| CAHFVNKLLNTHHDPEPF | TRBV28 | 5 | 7.25E-31 |
| CGGGARKTGQSPRQPRPGPSF | TRBV22-1 | 13 | 9.43E-32 |
| CGSTFEAGHGGGPDGYVVGRF | TRBV8-2 | 16 | 1.74E-33 |
| CGSTFEAGHGGGPDGYVMGRF | TRBV8-2 | 3 | 4.82E-34 |
| CECAGHGLWTCQEQQSRAPAF | TRBV5-2 | 2 | 6.91E-37 |
| CNDVAIFCVDLGYGANIPDHTQYF | TRBV4-1 | 8 | 5.31E-37 |
| CSGLCSSRTGNWAGAPGRASTGCSF | TRBV2 | 2 | 2.71E-37 |
| CGLCDMNMLCRHSLLFIFEFTF | TRBV5-2 | 2 | 2.07E-39 |

**Table S5**
$P_{gen}$ values for the $TCR\alpha^{aa}$ clonotypes that were shared between TCLs.

| **Predictive CDR3 amino acid sequence** | ***TRAV* genes** | **Number of shared samples** | **$P_{gen}$ value** |
|---|---|---|---|
| CAVTGNQFYF | TRAV41 | 3 | 2.50E-05 |
| CASGGSYIPTF | TRAV13-1 | 2 | 1.29E-05 |
| CAATDSWGKLQF | TRAV13-1 | 4 | 9.43E-06 |
| CAVMDSNYQLIW | TRAV1-2 | 4 | 9.13E-06 |
| CAYSGAGSYQLTF | TRAV27 | 2 | 5.06E-06 |
| CVVSSGGYNKLIF | TRAV10 | 3 | 5.04E-06 |
| CAGRLIQGAQKLVF | TRAV20 | 2 | 1.01E-06 |
| CDNNNDMRF | TRAV16 | 213 | 9.38E-07 |
| CAVRVGQKLLF | TRAV1-1 | 2 | 5.17E-07 |
| CALSGVTSGTYKYIF | TRAV19 | 2 | 2.25E-07 |
| CIASNAGGTSYGKLTF | TRAV26-1 | 2 | 2.02E-07 |
| CELSF | TRAV13-1 | 2 | 1.61E-07 |
| CAPIIF | TRAV17 | 4 | 1.19E-07 |
| WLSF | TRAV3 | 5 | 5.96E-08 |
| CALFF | TRDV1 | 12 | 2.69E-08 |
| CVVF | TRAV10 | 9 | 1.48E-08 |
| CAFMKVSSGTYKYIF | TRAV38-1 | 2 | 1.44E-08 |
| CLFF | TRAV17 | 2 | 9.81E-09 |
| CCGNNNARLMF | TRAV28, | 26 | 7.81E-09 |
| CALTF | TRAV33 | 3 | 7.33E-09 |
| CVVGLFF | TRAV10 | 3 | 2.62E-09 |
| CGCENSGGSNYKLTF | TRAV3 | 129 | 1.56E-09 |
| CCLIF | TRAV31 | 2 | 7.34E-10 |
| CAFMGLFF | TRAV38-1 | 2 | 2.78E-10 |
| CATAGQYNAGNMLTF | TRAV17 | 3 | 1.69E-10 |
| WFWLIF | TRAV34 | 24 | 8.05E-11 |

| | | | |
|---|---|---|---|
| CCGIPLVF | TRAV28 | 2 | 6.89E-12 |
| CALDSQPLIW | TRAV6 | 21 | 1.26E-12 |
| WQELLAF | TRAV27 | 5 | 7.35E-13 |
| CCYKELLF | TRAV31 | 5 | 4.06E-13 |
| SNGHLVF | TRAV1-2 | 2 | 1.19E-13 |
| FLQRYLFF | TRAV40 | 16 | 9.38E-14 |
| CESLKPLFF | TRAV16 | 2 | 2.07E-14 |
| CAENVYNQVGKLIF | TRAV5 | 2 | 1.58E-14 |
| CSLTCQSYKLSF | TRAV11 | 12 | 7.62E-16 |
| CYEASHSVNTGTASKLTF | TRAV31 | 64 | 3.31E-17 |
| CSRYYRAPELIF | TRAV10 | 97 | 1.33E-18 |
| CEVKDMFYLFF | TRAV2 | 3 | 7.83E-19 |
| CAAGELGGTASRF | TRAV8-5 | 2 | 4.60E-19 |
| CERNTDRNLVF | TRAV13-2 | 5 | 4.28E-19 |
| LLWKASSLKLIW | TRAV22 | 14 | 2.01E-20 |
| CAPVQRHRKYVVF | TRAV41 | 2 | 1.21E-21 |
| CDINFPEAITLIF | TRAV20 | 13 | 5.74E-22 |
| CEVENIQARPLFF | TRAV36DV7 | 4 | 1.26E-22 |
| CASRYSDVWIALIW | TRAV15 | 32 | 3.43E-23 |
| CCVWHYVDLSLVF | TRAV31 | 2 | 3.83E-24 |
| CISTSTSSTSTPIPTF | TRAV41 | 2 | 2.54E-24 |
| WCVWHYVDLSLVF | TRAV31 | 2 | 1.15E-24 |
| FAEMFEHFSLFF | TRAV5 | 19 | 7.46E-25 |
| CLDFVKIKVVIWLIF | TRAV38-1 | 2 | 2.48E-25 |
| CSYHNRQTSRKNLFF | TRAV26-2 | 5 | 1.26E-25 |
| CACDPGWSFTLLHFYF | TRDV2 | 2 | 7.63E-26 |
| CACDPGWSLMLLHFYF | TRDV2 | 3 | 2.84E-26 |

**Table S6**
$P_{gen}$ values for the most frequent TCRβ clonotype in TCLs.

| Predictive CDR3 amino acid sequence | *TRBV* genes | *TRBJ* genes | $P_{gen}$ value |
|---|---|---|---|
| CASSLAGTDTQYF | TRBV7-2 | TRBJ2-3 | 2.33E-06 |
| CASSLGGEQYF | TRBV7-7 | TRBJ2-7 | 2.27E-06 |
| CASSLTGNTEAFF | TRBV7-9 | TRBJ1-1 | 1.17E-06 |
| CASSEGGYGYTF | TRBV6-1 | TRBJ1-2 | 8.72E-07 |
| CASSPSYNEQFF | TRBV3-1 | TRBJ2-1 | 8.46E-07 |
| CASSYRGTDTQYF | TRBV6-6 | TRBJ2-3 | 7.27E-07 |
| CASSGTNYGYTF | TRBV19 | TRBJ1-2 | 7.26E-07 |
| CASSDYNEQFF | TRBV7-9 | TRBJ2-1 | 7.03E-07 |
| CASSLLAGGTDTQYF | TRBV7-9 | TRBJ2-3 | 4.23E-07 |
| CASSLDGSYEQYF | TRBV11-3 | TRBJ2-7 | 3.96E-07 |
| CASSSASSYEQYF | TRBV19 | TRBJ2-7 | 2.75E-07 |
| CASSDSSTDTQYF | TRBV6-4 | TRBJ2-3 | 2.69E-07 |
| CASSSGANVLTF | TRBV5-1 | TRBJ2-6 | 2.64E-07 |
| CASSLGGGANVLTF | TRBV7-9 | TRBJ2-6 | 2.51E-07 |
| CASSTGQGTEAFF | TRBV11-3 | TRBJ1-1 | 2.38E-07 |
| CASSKQGGYTF | TRBV6-1 | TRBJ1-2 | 2.23E-07 |
| CASSLAGTGGYEQYF | TRBV5-1 | TRBJ2-7 | 2.11E-07 |
| CASRQGAGELFF | TRBV11-3 | TRBJ2-2 | 1.81E-07 |
| CASSETGDYGYTF | TRBV6-6 | TRBJ1-2 | 1.81E-07 |
| CASSD*RSTDTQYF | TRBV25-1 | TRBJ2-3 | 1.67E-07 |
| CSARTGGYGYTF | TRBV20-1 | TRBJ1-2 | 1.67E-07 |
| CASSYTGNYGYTF | TRBV6-5 | TRBJ1-2 | 1.45E-07 |
| CASSLQDTGELFF | TRBV19 | TRBJ2-2 | 1.45E-07 |
| CASSYGTNQPQHF | TRBV6-5 | TRBJ1-5 | 1.36E-07 |
| CSASRSNQPQHF | TRBV20-1 | TRBJ1-5 | 1.21E-07 |
| CASSLARGSPLHF | TRBV5-6 | TRBJ1-6 | 1.14E-07 |

| | | | |
|---|---|---|---|
| CASSIDRGYGYTF | TRBV19 | TRBJ1-2 | 1.14E-07 |
| CASSQSGANVLTF | TRBV3-1 | TRBJ2-6 | 1.08E-07 |
| CASSPRTGGSNQPQHF | TRBV19 | TRBJ1-5 | 9.27E-08 |
| CASSRASGTDTQYF | TRBV4-1 | TRBJ2-3 | 8.82E-08 |
| CASGRGGGYTF | TRBV30 | TRBJ1-2 | 8.52E-08 |
| CASSHYTDTQYF | TRBV7-2 | TRBJ2-3 | 8.48E-08 |
| CASSLPSGTEAFF | TRBV5-6 | TRBJ1-1 | 8.23E-08 |
| CASSRGTSGSYNEQFF | TRBV12-3 | TRBJ2-1 | 8.19E-08 |
| CASSIGQGTDTQYF | TRBV19 | TRBJ2-3 | 7.11E-08 |
| CASSPGLAGGTYEQYF | TRBV7-2 | TRBJ2-7 | 5.45E-08 |
| CASSEVRGTEAFF | TRBV6-1 | TRBJ1-1 | 5.20E-08 |
| CSARGQNTGELFF | TRBV20-1 | TRBJ2-2 | 5.18E-08 |
| CASSYSTGEAFF | TRBV6-5 | TRBJ1-1 | 4.90E-08 |
| CASRDIQETQYF | TRBV10-3 | TRBJ2-5 | 4.85E-08 |
| CASSIGRADTQYF | TRBV19 | TRBJ2-3 | 4.82E-08 |
| CASSLESGSYNEQFF | TRBV11-3 | TRBJ2-1 | 3.40E-08 |
| CASSLGTGGLQPQHF | TRBV28 | TRBJ1-5 | 2.37E-08 |
| CASSYSTGLSYEQYF | TRBV6-6 | TRBJ2-7 | 2.20E-08 |
| CASSVDTGNQPQHF | TRBV9 | TRBJ1-5 | 2.13E-08 |
| CASSYSGTGRYEQYF | TRBV6-5 | TRBJ2-7 | 2.09E-08 |
| CASNPTGSNQPQHF | TRBV6-1 | TRBJ1-5 | 2.02E-08 |
| CASSLQGSGQPQHF | TRBV27 | TRBJ1-5 | 1.74E-08 |
| CASSLNQDTEAFF | TRBV11-3 | TRBJ1-1 | 1.72E-08 |
| CASSESGQTYEQYF | TRBV10-2 | TRBJ2-7 | 1.71E-08 |
| CSVEAFF | TRBV29-1 | TRBJ1-1 | 1.57E-08 |
| CASSLGDGYNSPLHF | TRBV5-6 | TRBJ1-6 | 1.37E-08 |
| CATSDLLAGEETQYF | TRBV24-1 | TRBJ2-5 | 1.25E-08 |
| CSARDGLAGDTQYF | TRBV20-1 | TRBJ2-3 | 1.24E-08 |
| CSASGSRETQYF | TRBV20-1 | TRBJ2-5 | 1.21E-08 |
| CAWSVQGDTQYF | TRBV30 | TRBJ2-3 | 1.11E-08 |
| CASSSLTGTYGYTF | TRBV6-5 | TRBJ1-2 | 1.03E-08 |

| | | | |
|---|---|---|---|
| CSARRGGRTEAFF | TRBV20-1 | TRBJ1-1 | 1.01E-08 |
| CASSFIPGTDTQYF | TRBV5-6 | TRBJ2-3 | 9.56E-09 |
| CASSYSI*GYEQYF | TRBV6-6 | TRBJ2-7 | 9.11E-09 |
| CATSERQSNQPQHF | TRBV24-1 | TRBJ1-5 | 8.24E-09 |
| CASSEDGGSPLHF | TRBV6-1 | TRBJ1-6 | 7.81E-09 |
| CASSLETEGTEAFF | TRBV5-1 | TRBJ1-1 | 7.74E-09 |
| CSARRDSSSYEQYF | TRBV20-1 | TRBJ2-7 | 7.66E-09 |
| CASSPLDRGGEKLFF | TRBV3-1 | TRBJ1-4 | 7.27E-09 |
| CSATGTGTNEKLFF | TRBV20-1 | TRBJ1-4 | 7.26E-09 |
| CASSPRLAGVSSYNEQFF | TRBV7-9 | TRBJ2-1 | 7.11E-09 |
| CASSWRTGDEKLFF | TRBV28 | TRBJ1-4 | 6.46E-09 |
| CASSFRLEPYEQYF | TRBV5-6 | TRBJ2-7 | 6.02E-09 |
| CASSLD*REETDTQYF | TRBV5-6 | TRBJ2-3 | 6.01E-09 |
| CATSRGGHQETQYF | TRBV24-1 | TRBJ2-5 | 5.91E-09 |
| CASSDNRGSQPQHF | TRBV6-1 | TRBJ1-5 | 4.99E-09 |
| CASHPGSSYNEQFF | TRBV5-6 | TRBJ2-1 | 4.92E-09 |
| CASGAGVGTDTQYF | TRBV5-1 | TRBJ2-3 | 4.72E-09 |
| CASSVEGFSYNEQFF | TRBV9 | TRBJ2-1 | 4.37E-09 |
| CASSKVTGGTDTQYF | TRBV10-2 | TRBJ2-3 | 3.91E-09 |
| CASSAETGGGYEQYF | TRBV6-1 | TRBJ2-7 | 3.49E-09 |
| CASSSGLAAPTDTQYF | TRBV7-3 | TRBJ2-3 | 3.08E-09 |
| CASSYSGLARNEQFF | TRBV6-5 | TRBJ2-1 | 2.67E-09 |
| CSASAGADGYTF | TRBV20-1 | TRBJ1-2 | 2.53E-09 |
| CSVGPQGTDTQYF | TRBV29-1 | TRBJ2-3 | 2.49E-09 |
| CASSQVAGGVYNEQFF | TRBV4-1 | TRBJ2-1 | 2.35E-09 |
| CASKRGTSETQYF | TRBV19 | TRBJ2-5 | 2.33E-09 |
| CSAVAGFSYEQYF | TRBV20-1 | TRBJ2-7 | 2.30E-09 |
| CASSYTGSGAYEQYF | TRBV6-2 | TRBJ2-7 | 2.28E-09 |
| CASSDAGGRSPLHF | TRBV6-5 | TRBJ1-6 | 2.12E-09 |
| CASSISGGEQPQHF | TRBV19 | TRBJ1-5 | 1.98E-09 |
| CSASAGINYGYTF | TRBV20-1 | TRBJ1-2 | 1.89E-09 |

| | | | |
|---|---|---|---|
| CSVSPLETQYF | TRBV20-1 | TRBJ2-5 | 1.63E-09 |
| CASSYTSGPNQPQHF | TRBV6-5 | TRBJ1-5 | 1.53E-09 |
| CASRLGTGGGQETQYF | TRBV5-1 | TRBJ2-5 | 1.41E-09 |
| CASSEQESSPLHF | TRBV6-4 | TRBJ1-6 | 1.30E-09 |
| CASSLGVAGEGYEQYF | TRBV11-3 | TRBJ2-7 | 1.18E-09 |
| CSVEGQGVSGANVLTF | TRBV29-1 | TRBJ2-6 | 1.17E-09 |
| CAWSVGVTYGYTF | TRBV30 | TRBJ1-2 | 1.10E-09 |
| CSATQGQDTQYF | TRBV20-1 | TRBJ2-3 | 1.10E-09 |
| CASILGPSNTGELFF | TRBV7-2 | TRBJ2-2 | 9.64E-10 |
| CASSLGTPLANTGELFF | TRBV27 | TRBJ2-2 | 6.09E-10 |
| CASSPRGSGTEQFF | TRBV11-3 | TRBJ2-1 | 5.07E-10 |
| CSARLDGYYGYTF | TRBV20-1 | TRBJ1-2 | 5.05E-10 |
| CASSSLKGDSYNEQFF | TRBV5-4 | TRBJ2-1 | 4.74E-10 |
| CASSYSKGLVSYNEQFF | TRBV6-2 | TRBJ2-1 | 4.73E-10 |
| CASSSQQGGLTEAFF | TRBV7-2 | TRBJ1-1 | 4.69E-10 |
| CASSEGTTGNNEQFF | TRBV6-1 | TRBJ2-1 | 4.46E-10 |
| CASSLQAPRSGNTIYF | TRBV7-8 | TRBJ1-3 | 3.67E-10 |
| CASSDTRTGSNTGELFF | TRBV9 | TRBJ2-2 | 2.95E-10 |
| CSARERRDPSYEQYF | TRBV20-1 | TRBJ2-7 | 2.33E-10 |
| CSAERHHNEQFF | TRBV20-1 | TRBJ2-1 | 2.22E-10 |
| CSALGGGGIEQYF | TRBV20-1 | TRBJ2-7 | 2.11E-10 |
| CASSQRQLGGETQYF | TRBV5-6 | TRBJ2-5 | 1.95E-10 |
| CASSIGQGAQSPLHF | TRBV3-1 | TRBJ1-6 | 1.92E-10 |
| CSARVASVSTDTQYF | TRBV20-1 | TRBJ2-3 | 1.83E-10 |
| CASNYEGANTEAFF | TRBV5-1 | TRBJ1-1 | 1.34E-10 |
| CSAPETGGSGYTF | TRBV20-1 | TRBJ1-2 | 1.30E-10 |
| CASRPGTQNNSPLHF | TRBV19 | TRBJ1-6 | 9.39E-11 |
| CASSQVGRGGLEAFF | TRBV14 | TRBJ1-1 | 8.72E-11 |
| CASSPLVGRIQETQYF | TRBV5-1 | TRBJ2-5 | 8.57E-11 |
| CASSLGTSGRMSGELFF | TRBV5-6 | TRBJ2-2 | 7.38E-11 |
| CASSLEEGAGFGYTF | TRBV5-6 | TRBJ1-2 | 5.97E-11 |

| | | | |
|---|---|---|---|
| CASSRPWGGFDEQYF | TRBV18 | TRBJ2-7 | 4.90E-11 |
| CSARSASGGAGTDTQYF | TRBV20-1 | TRBJ2-3 | 4.73E-11 |
| CASTQRGDGTYEQYF | TRBV19 | TRBJ2-7 | 3.33E-11 |
| CASSQAGTDTQFF | TRBV4-2 | TRBJ2-3 | 2.90E-11 |
| CASSYRDKFRGYTF | TRBV6-6 | TRBJ1-2 | 2.89E-11 |
| CASSQAGGLLLTYEQYF | TRBV11-3 | TRBJ2-7 | 1.94E-11 |
| CASSKFGLCSGANVLTF | TRBV19 | TRBJ2-6 | 1.92E-11 |
| CSASLLGGVSNYGYTF | TRBV20-1 | TRBJ1-2 | 1.22E-11 |
| CASSVGESVFGYTF | TRBV6-1 | TRBJ1-2 | 1.13E-11 |
| CASSQQIGQKNTEAFF | TRBV4-1 | TRBJ1-1 | 1.07E-11 |
| CASAPTAWDTQYF | TRBV6-1 | TRBJ2-3 | 8.85E-12 |
| CASQRRGSRDTEAFF | TRBV27 | TRBJ1-1 | 8.79E-12 |
| CASCTPGQGARYEQYF | TRBV6-1 | TRBJ2-7 | 8.54E-12 |
| CASSVVGRVDAEAFF | TRBV9 | TRBJ1-1 | 8.32E-12 |
| CASSFEPTSGGANEQYF | TRBV7-8 | TRBJ2-7 | 8.23E-12 |
| CAWGEGQGFETQYF | TRBV30 | TRBJ2-5 | 7.03E-12 |
| WSCLF | TRBV22-1 | TRBJ2-2 | 6.64E-12 |
| CSVSGTGQLAYEQYF | TRBV29-1 | TRBJ2-7 | 5.92E-12 |
| CASSYYTGRYNTGELFF | TRBV6-5 | TRBJ2-2 | 5.47E-12 |
| CASSIVGVTSYSNQPQHF | TRBV19 | TRBJ1-5 | 4.98E-12 |
| CASSTRAGGSTSSYNEQFF | TRBV19 | TRBJ2-1 | 3.91E-12 |
| CSAKRGLAGVGGELFF | TRBV20-1 | TRBJ2-2 | 3.37E-12 |
| CASNPEVHKETQYF | TRBV5-4 | TRBJ2-5 | 3.00E-12 |
| CASSGMEGQGALYGYTF | TRBV28 | TRBJ1-2 | 2.24E-12 |
| CASRPSGDPPPLHF | TRBV5-4 | TRBJ1-6 | 1.94E-12 |
| CASRLAGATHRQYF | TRBV3-1 | TRBJ2-3 | 4.19E-13 |
| CATSDTPGQEFFF | TRBV24-1 | TRBJ1-1 | 1.38E-13 |
| CSIEDGAVYGYTF | TRBV29-1 | TRBJ1-2 | 9.78E-14 |
| CASSQTHWLAVHEQYF | TRBV23-1 | TRBJ2-7 | 3.07E-14 |
| CSVEDVIYDYTF | TRBV29-1 | TRBJ1-2 | 2.04E-14 |
| CASSIVTGNYAYTF | TRBV19 | TRBJ1-2 | 1.47E-14 |

| | | | |
|---|---|---|---|
| CASSLDHIDSGGTPYEQYF | TRBV6-5 | TRBJ2-7 | 6.62E-15 |
| CSARARGDSDSPSSYNEQFF | TRBV20-1 | TRBJ2-1 | 4.76E-15 |
| CATSRLIPGLAGVGEAKNIQYF | TRBV15 | TRBJ2-4 | 2.14E-15 |
| CARSISGQGAGGSHEQYF | TRBV10-1 | TRBJ2-7 | 3.92E-16 |
| CAK*KPRFRGLF | TRBV6-4 | TRBJ1-1 | 1.42E-16 |
| CASSIEMTTNSNHPQHF | TRBV6-6 | TRBJ1-5 | 9.59E-20 |
| CASSEGLAADNRVYDKQYF | TRBV6-1 | TRBJ2-7 | 1.15E-20 |
| CASSAQGVKMPGGGWEDTEAFF | TRBV7-9 | TRBJ1-1 | 6.93E-21 |
| CDISQHYDHGFQYF | TRBV10-3 | TRBJ2-4 | 8.58E-23 |
| WHPSHLLFFFFVF | TRBV6-2 | TRBJ2-2 | 4.89E-24 |
| CAWSPTGPKMRSGFCAGLGGRDQETQYF | TRBV30 | TRBJ2-5 | 2.60E-31 |
| SGQQLLKRRMPFLPVCESVLTF | TRBV7-4 | TRBJ2-6 | 2.86E-36 |
| CAKQLNEDCSKTQPCDHTKGLECNF | TRBV23-1 | TRBJ2-4 | 5.16E-44 |

**Table S7**
$P_{gen}$ values for the most frequent TCRα clonotype in samples of subgroups of TCLs.

| Predictive CDR3 amino acid sequence | *TRAV* genes | *TRAJ* genes | $P_{gen}$ value |
|---|---|---|---|
| CAATGNQFYF | TRAV29DV5 | TRAJ49 | 3.80E-05 |
| CAASTGNQFYF | TRAV23DV6 | TRAJ49 | 2.81E-05 |
| CAVTGNQFYF | TRAV41 | TRAJ49 | 2.50E-05 |
| CAASNDYKLSF | TRAV13-1 | TRAJ20 | 2.03E-05 |
| CAANQGGKLIF | TRAV13-1 | TRAJ23 | 1.44E-05 |
| CAASGGSSNTGKLIF | TRAV13-1 | TRAJ37 | 1.36E-05 |
| CAVSGSARQLTF | TRAV8-4 | TRAJ22 | 1.34E-05 |
| CAASSSASKIIF | TRAV13-1 | TRAJ3 | 1.27E-05 |
| CAVFSGGYNKLIF | TRAV1-2 | TRAJ4 | 1.27E-05 |
| CAGDTGRRALTF | TRAV25 | TRAJ5 | 1.25E-05 |
| CALRYSSASKIIF | TRAV9-2 | TRAJ3 | 9.76E-06 |
| CAATTSGTYKYIF | TRAV21 | TRAJ40 | 9.67E-06 |

| | | | |
|---|---|---|---|
| CAVMDSNYQLIW | TRAV1-2 | TRAJ33 | 9.13E-06 |
| CAGDMRF | TRAV20 | TRAJ43 | 7.89E-06 |
| CAASIKAAGNKLTF | TRAV13-1 | TRAJ17 | 7.87E-06 |
| CAVIYNQGGKLIF | TRAV12-2 | TRAJ23 | 6.99E-06 |
| CAVNSGGYQKVTF | TRAV1-2 | TRAJ13 | 6.33E-06 |
| CAVGDYKLSF | TRAV8-3 | TRAJ20 | 6.03E-06 |
| CAERFSDGQKLLF | TRAV13-2 | TRAJ16 | 5.61E-06 |
| CAVGQGAQKLVF | TRAV41 | TRAJ54 | 4.72E-06 |
| CAAIFGNEKLTF | TRAV29DV5 | TRAJ48 | 4.21E-06 |
| CAVAGSARQLTF | TRAV8-3 | TRAJ22 | 3.76E-06 |
| CAVSAAGNKLTF | TRAV1-1 | TRAJ17 | 3.53E-06 |
| CAVGDDKIIF | TRAV8-3 | TRAJ30 | 3.39E-06 |
| CAGMNTGFQKLVF | TRAV8-3 | TRAJ8 | 3.23E-06 |
| CAMREGSSNTGKLIF | TRAV14DV4 | TRAJ37 | 3.05E-06 |
| CAARGANSKLTF | TRAV13-1 | TRAJ56 | 2.89E-06 |
| CAASRGSGGSNYKLTF | TRAV13-1 | TRAJ53 | 2.49E-06 |
| CAVRDQAGTALIF | TRAV3 | TRAJ15 | 2.45E-06 |
| CALSDAGNNRKLIW | TRAV9-2 | TRAJ38 | 2.09E-06 |
| CAVRRSGGSYIPTF | TRAV21 | TRAJ6 | 1.70E-06 |
| CALRGNTGKLIF | TRAV19 | TRAJ37 | 1.68E-06 |
| CATGSGNTPLVF | TRAV17 | TRAJ29 | 1.66E-06 |
| CAGTSGNTPLVF | TRAV25 | TRAJ29 | 1.64E-06 |
| CNTNAGKSTF | TRAV35 | TRAJ27 | 1.57E-06 |
| CALKNQGGKLIF | TRAV9-2 | TRAJ23 | 1.35E-06 |
| CAVNNYGQNFVF | TRAV21 | TRAJ26 | 1.33E-06 |
| CLTSGTYKYIF | TRAV17 | TRAJ40 | 1.03E-06 |
| CAASMGNTGNQFYF | TRAV13-1 | TRAJ49 | 1.02E-06 |
| CAESNNNARLMF | TRAV5 | TRAJ31 | 9.44E-07 |
| CDNNNDMRF | TRAV16 | TRAJ43 | 9.38E-07 |
| CAGIMDSNYQLIW | TRAV25 | TRAJ33 | 8.44E-07 |
| CALSFTGNQFYF | TRAV9-2 | TRAJ49 | 7.70E-07 |

| | | | |
|---|---|---|---|
| CAASHTSGTYKYIF | TRAV23DV6 | TRAJ40 | 7.61E-07 |
| CAARYSGNTPLVF | TRAV13-1 | TRAJ29 | 7.40E-07 |
| CAERDQTGANNLFF | TRAV13-2 | TRAJ36 | 6.59E-07 |
| CAEKGTGFQKLVF | TRAV5 | TRAJ8 | 5.96E-07 |
| CAASIRGGATNKLIF | TRAV13-1 | TRAJ32 | 5.86E-07 |
| CAVRVGQKLLF | TRAV1-1 | TRAJ16 | 5.17E-07 |
| CAARGDGGSQGNLIF | TRAV29DV5 | TRAJ42 | 4.22E-07 |
| CALGGGAQKLVF | TRAV9-2 | TRAJ54 | 4.20E-07 |
| CATDDNNNDMRF | TRAV17 | TRAJ43 | 4.04E-07 |
| CAIRNSGNTPLVF | TRAV12-3 | TRAJ29 | 3.47E-07 |
| CAAAGYNFNKFYF | TRAV13-1 | TRAJ21 | 3.21E-07 |
| CAGAIGSARQLTF | TRAV27 | TRAJ22 | 2.82E-07 |
| CAVQGGTASKLTF | TRAV13-1 | TRAJ44 | 2.53E-07 |
| CALIYNTNAGKSTF | TRAV9-2 | TRAJ27 | 2.45E-07 |
| CAESSQGGKLIF | TRAV5 | TRAJ23 | 2.32E-07 |
| CAASVGTALIF | TRAV13-1 | TRAJ15 | 2.24E-07 |
| CAMTRGSSNTGKLIF | TRAV14DV4 | TRAJ37 | 1.94E-07 |
| CIVRASGGYQKVTF | TRAV26-1 | TRAJ13 | 1.93E-07 |
| CAMSADKLIF | TRAV12-3 | TRAJ34 | 1.78E-07 |
| CAVSERDDKIIF | TRAV8-4 | TRAJ30 | 1.75E-07 |
| CLVGVRNSGNTPLVF | TRAV4 | TRAJ29 | 1.58E-07 |
| CAESINTNAGKSTF | TRAV5 | TRAJ27 | 1.50E-07 |
| CAYRSEGNKLVF | TRAV38-2DV8 | TRAJ47 | 1.46E-07 |
| CAASNDTDKLIF | TRAV13-1 | TRAJ34 | 1.37E-07 |
| CAMSLFSDGQKLLF | TRAV14DV4 | TRAJ16 | 1.24E-07 |
| CAYRSASGTSYGKLTF | TRAV38-2DV8 | TRAJ52 | 1.22E-07 |
| CAGPNYGGATNKLIF | TRAV27 | TRAJ32 | 1.01E-07 |
| CIVRVGPGGYQKVTF | TRAV26-1 | TRAJ13 | 9.16E-08 |
| CTYGSSNTGKLIF | TRAV38-2DV8 | TRAJ37 | 8.89E-08 |
| CAFMDAGNNRKLIW | TRAV38-1 | TRAJ38 | 7.24E-08 |
| CAASINGFQKLVF | TRAV13-1 | TRAJ8 | 6.23E-08 |

| | | | |
|---|---|---|---|
| CAVVAGQKLLF | TRAV2 | TRAJ16 | 4.47E-08 |
| CALSEQASGTYKYIF | TRAV19 | TRAJ40 | 4.09E-08 |
| CAVTLMIYNQGGKLIF | TRAV8-4 | TRAJ23 | 3.72E-08 |
| CAAMWGSSNTGKLIF | TRAV29DV5 | TRAJ37 | 3.60E-08 |
| CAVNRGGANNLFF | TRAV12-2 | TRAJ36 | 2.62E-08 |
| CAMSEGGGGNKLTF | TRAV12-3 | TRAJ10 | 2.30E-08 |
| CAISHSGGYQKVTF | TRAV9-2 | TRAJ13 | 2.17E-08 |
| CAMSAYNYGQNFVF | TRAV12-3 | TRAJ26 | 1.94E-08 |
| CAVIPLSGTYKYIF | TRAV8-1 | TRAJ40 | 1.87E-08 |
| CALNINAGKSTF | TRAV9-2 | TRAJ27 | 1.56E-08 |
| CAMR*NGSSNTGKLIF | TRAV12-3 | TRAJ37 | 1.50E-08 |
| CAFMKVSSGTYKYIF | TRAV38-1 | TRAJ40 | 1.44E-08 |
| CALSEWRTSGTYKYIF | TRAV19 | TRAJ40 | 9.91E-09 |
| CAVPFEGAQKLVF | TRAV41 | TRAJ54 | 8.27E-09 |
| CALSNVAGGTSYGKLTF | TRAV9-2 | TRAJ52 | 7.90E-09 |
| CATGGLYGNKLVF | TRAV17 | TRAJ47 | 7.21E-09 |
| CVVRRKLTGGGNKLTF | TRAV10 | TRAJ10 | 6.46E-09 |
| CAVGAQNARLMF | TRAV8-3 | TRAJ31 | 4.39E-09 |
| CGCENSGGSNYKLTF | TRAV3 | TRAJ53 | 1.56E-09 |
| CAASIQGGGGKLIF | TRAV13-1 | TRAJ23 | 1.13E-09 |
| CAFKSRGGSEKLVF | TRAV38-1 | TRAJ57 | 8.75E-10 |
| CVVRFPFNDYKLSF | TRAV1-2 | TRAJ20 | 1.91E-10 |
| CAMREGWGLAF | TRAV14DV4 | TRAJ7 | 1.86E-10 |
| CALGELGNARLMF | TRDV1 | TRAJ31 | 1.64E-10 |
| CIVKKRF | TRAV26-1 | TRAJ43 | 8.94E-11 |
| CDVTGAGNNRKLIW | TRAV12-2 | TRAJ38 | 4.10E-11 |
| CALSGTSETYKYIF | TRAV9-2 | TRAJ40 | 1.96E-11 |
| CIPRDVGNEKLTF | TRAV26-2 | TRAJ48 | 1.77E-11 |
| CALMKRGTNDMRF | TRAV38-1 | TRAJ43 | 1.40E-12 |
| CAVKFSGGCNKLIF | TRAV12-2 | TRAJ4 | 2.95E-15 |
| CALRRHRGIRSTDKLIF | TRDV1 | TRDJ1 | 1.68E-16 |

| | | | |
|---|---|---|---|
| CALGEISYVPSTDKLIF | TRDV1 | TRDJ1 | 5.89E-18 |
| SAGQLGTGKQFYF | TRAV35 | TRAJ49 | 2.85E-20 |
| CALSDHRDWGIRSRPLIF | TRAV19 | TRDJ4 | 2.94E-22 |
| CALGELRGINPTVLGGKLIF | TRDV1 | TRDJ1 | 1.20E-22 |
| CACDAIPRPTGAADKLIF | TRDV2 | TRDJ1 | 8.31E-23 |
| CACDTVSGGYANPDKLIF | TRDV2 | TRDJ1 | 9.77E-24 |

9. Wang L, Ni X, Covington KR, Yang BY, Shiu J, Zhang X, et al. Genomic profiling of Sézary syndrome identifies alterations of key T cell signaling and differentiation genes. Nature

Genetics. 2015. pp. 1426–1434. doi:10.1038/ng.3444

10. Yoo HY, Sung MK, Lee SH, Kim S, Lee H, Park S, et al. A recurrent inactivating mutation in RHOA GTPase in angioimmunoblastic T cell lymphoma. Nat Genet. 2014;46: 371–375.

11. Jiang L, Gu Z-H, Yan Z-X, Zhao X, Xie Y-Y, Zhang Z-G, et al. Exome sequencing identifies somatic mutations of DDX3X in natural killer/T-cell lymphoma. Nat Genet. 2015;47: 1061–1066.

12. Crescenzo R, Abate F, Lasorsa E, Tabbo' F, Gaudiano M, Chiesa N, et al. Convergent mutations and kinase fusions lead to oncogenic STAT3 activation in anaplastic large cell lymphoma. Cancer Cell. 2015;27: 516–532.

13. Kataoka K, Nagata Y, Kitanaka A, Shiraishi Y, Shimamura T, Yasunaga J-I, et al. Integrated molecular analysis of adult T cell leukemia/lymphoma. Nat Genet. 2015;47: 1304–1315.
